# Beta-blockers or Placebo for Primary Prophylaxis (BOPPP) of oesophageal varices: Study protocol for a randomised controlled trial

**DOI:** 10.1101/2023.12.21.23300362

**Authors:** Vishal C Patel, Mark J McPhail, Ruhama Uddin, Hassan Jafari, Vanessa Lawrence, Clair Le Boutillier, James Shearer, Nahel Yaziji, Angela Cape, Haroon Ahmed, Christopher Ward, Peter Walsh, Kevin Besly, Ane Zamalloa, Joanna Kelly, BOPPP study group, Ben Carter

## Abstract

**Background:** Liver disease is within the top five causes of premature death in adults. Deaths caused by complications of cirrhosis continue to rise, while deaths related to other non-liver disease areas are declining. Portal hypertension, the primary sequalae of cirrhosis and is associated with the development of variceal haemorrhage, ascites, hepatic encephalopathy and infection, collectively termed hepatic decompensation, that lead to hospitalisation and mortality. It remains uncertain whether administering a non-selective beta-blocker (NSBB), specifically carvedilol, at an earlier stage i.e. when oesophageal varices are small, can prevent VH and reduce all-cause decompensation (ACD).

**Methods/design:** The BOPPP trial is a pragmatic, multicentre, placebo-controlled, triple-blinded, randomised controlled trial (RCT) in England, Scotland, Wales and Northern Ireland. Patients aged 18 years or older with cirrhosis and small oesophageal varices that have never bled will be recruited, subject to exclusion criteria. The trial aims to enrol 740 patients across 55 hospitals in the UK. Patients are allocated randomly on a 1:1 ratio to receive either carvedilol 6.25mg (a NSBB) or a matched placebo, once or twice daily, for 36 months, to attain adequate power to determine the effectiveness of carvedilol in preventing or reducing ACD.

The primary outcome is time to first decompensating event. It is a composite primary outcome made up of variceal haemorrhage (VH, new or worsening ascites, new or worsening hepatic encephalopathy (HE), spontaneous bacterial peritonitis (SBP), hepatorenal syndrome, an increase in Child Pugh grade by 1 grade or MELD score by 5 points, and liver-related mortality. Secondary outcomes include progression to medium or large oesophageal varices, development of gastric, duodenal, or ectopic varices, participant quality of life, healthcare costs and transplant-free survival.

**Discussion:** The BOPPP trial aims to investigate the clinical and cost effectiveness of carvedilol in patients with cirrhosis and small oesophageal varices to determine whether this non-selective beta-blocker can prevent or reduce hepatic decompensation. There is clinical equipoise on whether intervening in cirrhosis, at an earlier stage of portal hypertension, with NSBB therapy is beneficial. Should the trial yield a positive result, we anticipate that the administration and use of carvedilol will become widespread with pathways developed to standardise the administration of the medication in primary care.

**Ethics and dissemination:** The trial has been approved by the National Health Service (NHS) Research Ethics Committee (REC) (reference number: 19/YH/0015). The results of the trial will be submitted for publication in a peer-reviewed scientific journal. Participants will be informed of the results *via* the BOPPP website (www.boppp-trial.org) and partners in the British Liver Trust (BLT) organisation.

**Trial registration:** EUDRACT reference number: 2018-002509-78

ISRCTN reference number: ISRCTN10324656

## Introduction

### Background and trial rationale

Deaths due to complications of cirrhosis continue to rise while mortality rates from non-liver diseases are declining due to medical advances. In the UK, a 400% increase in mortality has been reported over a 40-year period from 1970 to 2010 in those with cirrhosis amongst those under 65 years old(1). Portal hypertension is the main complication of cirrhosis, which leads to the development of varices and variceal haemorrhage, and other forms of decompensation such as encephalopathy, ascites and renal failure(2). Patients with cirrhosis experience significant morbidity and reduced life expectancy due to all-cause decompensation(3). Despite therapeutic advances, the mortality rate for acute variceal haemorrhage (VH) remains approximately 15% (4). Currently, there are no established preventative methods (5), and thus, it is crucial to prevent VH and all-cause decompensation (ACD) in individuals who have developed varices as a manifestation of clinically-significant portal hypertension (CSPH).

Non-selective beta-blockade (NSBB) is the primary pharmacological choice and following several randomised controlled trials on various endoscopic methods, band ligation is now the preferred endoscopic therapy for medium and large oesophageal varices (OV)(6). NSBBs offer significant advantages by modulating portal hypertension, including reducing both the rate of incidence of primary and secondary VH, and progression of medium to larger varices(7). According to the current evidence base, NSBB therapy has no benefits in pre-primary prophylaxis for patients without varices, in part because large scale clinical trials to definitively address this are lacking(6). However, there is a clear advantage in the reduction of VH with NSBB in patients with moderate-large varices (>5mm in diameter), in patients with advanced cirrhosis(8, 9). There is currently no definitive evidence to provide clinical guidance on the use of NSBB in cirrhosis patients with compensated cirrhosis and/or small oesophageal varices(10), which are considered to be the precursor to development of medium or large varices as a reflection of escalating portal pressures. Moreover, it is unknown whether patients with small varices require primary prophylaxis at all.

The BOPPP trial aims to determine the clinical efficacy and cost effectiveness of NSBB therapy in patients with small oesophageal varices. NSBBs are low cost and easy to administer, making them suitable for primary healthcare settings. Due to their mechanism of action, by decreasing portal pressure which is the primary driver for ACD complications, NSBBs may also prevent the development of ascites, hepatic encephalopathy, hepatorenal syndrome, infections and liver synthetic failure. The international consensus in portal hypertension trials supports using of all-cause decompensation as the optimal endpoint.(11) All-cause decompensation encompasses progression of cirrhosis into clinical events such as development of ascites, hepatic encephalopathy, VH, hepatorenal syndrome and liver-related death(12). Several smaller trials in patients with invasive portal pressure measurements defining CSPH have indicated that NSBBs may have potential effects over some aspects of decompensation(9, 13), but this remains unproven in patients with small varices.

### Primary objective

The primary aim of this study is to determine the clinical effectiveness of carvedilol versus placebo in reducing all-cause decompensation in patients with cirrhosis and small oesophageal varices that have never bled. The study will also investigate the cost-effectiveness of administering carvedilol to these patients. All-cause decompensation (ACD) in the context of the BOPPP trial is defined in Table 1.

**Table 1.** Definition of ACD in the context of the BOPPP trial

| **Definition of composite primary outcome (All-cause decompensation)** |
| --- |
| • Variceal haemorrhage (VH)<br>• New or worsening ascites<br>• New or worsening hepatic encephalopathy<br>• Spontaneous bacterial peritonitis<br>• Hepatorenal syndrome<br>• Increase in Child Pugh Grade by 1 grade or increase MELD score by 5 points<br>• Liver-related mortality |

### Secondary objectives

- At 1-year after participant recruitment commences, to assess feasibility of recruitment and retention acceptability, with progression criteria outlined in the internal pilot.
- To determine additional clinical benefits of carvedilol versus placebo for reduction of variceal size progression, need to initiate endoscopic management of varices (endoscopic band ligation), deterioration in liver synthetic function (assessed by MELD score and Child-Pugh grade) and all-cause mortality.
- To determine the optimal delivery in primary care by exploring general practitioners’ (GPs) perspectives on enablers and barriers to future implementation

#### Rationale for changing primary outcome

The primary end point for BOPPP was initially variceal haemorrhage, as originally commissioned by the funder. This was based on 2017 national and international guidelines on cirrhosis that recommended that all patients with cirrhosis be offered surveillance for oesophageal varices. The 2015 British Society of Gastroenterology UK guidelines on the management of variceal haemorrhage in cirrhosis patients and other guidelines suggest that NSBB may be used as primary prophylaxis to prevent or reduce variceal bleeding in those identified as having small varices (<5mm)(14) [. However, they acknowledge that the evidence for this was weak and that further research is required to answer this long standing uncertainty. The NICE guideline “Cirrhosis in over 16s: assessment and management” [NG50] was published in July 2016(15) The evidence updates for this guideline confirm that the evidence on which to base recommendations for use of NSBBs for small varices is limited and warrants further research. Subsequent ‘Joint BSG/BASL Updated Guidance on Endoscopy for Variceal Screening and Surveillance in Chronic Liver Disease Patients in the COVID-19 Pandemic Service Recovery Phase’ acknowledged that COVID-19 pandemic led to many changes to endoscopic practice, including in patients with cirrhosis and recognised the need for ongoing research into the role of endoscopy and optimal management of portal hypertension in patients with chronic liver disease. The statement also acknowledged that NICE guidance in this field was due to be updated, the publication of a new consensus statement from Baveno VII (12), new UK guidelines in cirrhosis management in progress, and other work being done in the UK Gastroenterology and Hepatology community in this field currently, all of which mean that this interim guidance would be reviewed at a minimum interval of 12 months.

Note was made of Baveno VII that includes a consensus statement (statement 5.14) that treatment with NSBB should be considered for the prevention of decompensation in cirrhosis patients with clinically significant portal hypertension which is defined as Liver Stiffness Measurement (LSM) >25kPa (statement 2.16). This was acknowledged as being a controversial recommendation, being based on a single prospective trial and retrospective cohort evidence. The use of beta-blockers to prevent any decompensation episode was not in widespread hepatology practice in the UK at the time and there remains ongoing uncertainty regarding the PREDESCI data (13) when applying non-invasive tests to guide treatment in patient populations that are no longer seen in UK practice. In addition, BOPPP was highlighted as one of the ongoing large UK trials that would provide evidence in relation to the use of NSBB in those with cirrhosis and small varices. Therefore, no recommendation was given on the use of NSBB in cirrhosis patients with small varices but that this would remain under review.

Following recommendation and approval from trial oversight committees (TMG, DMC and TSC), PPI members and the funder, an amendment was submitted to change the primary outcome from variceal haemorrhage to a composite primary endpoint, all-cause decompensation (ACD). This modification of the primary outcome was motivated by (i) escalating evidence of the importance of this combined endpoint and (ii) a shift in scientific opinion from the global hepatology community. The definition of ACD as an endpoint has been ratified at the British Association for the Study of the Liver Research Steering Group. Furthermore, ACD, holds greater meaning and significance for patients with cirrhosis, as any worsening of their liver disease or need for hospitalisation due to deterioration of liver health is closely associated with patient’s quality of life and overall survival.

### Internal pilot

The first 12 months of BOPPP constituted an internal pilot. Integrated qualitative research with patient participants and staff contributed to assessments of feasibility and overall trial acceptability. The conclusions from the qualitative research led to positive changes in the trial protocol which maximised recruitment and reduced barriers that were identified to participation and retention. The results of the internal pilot were assessed by the TMG, DMC, TSC and the NIHR.

### Trial design

BOPPP is a phase IV, multicentre, blinded (patient, physician, analyst), randomised, placebo-controlled, pragmatic, clinical trial. The trial design is displayed in Figure 1. Patients with cirrhosis and small oesophageal varices receiving care from hospital specialist liver services are screened using the inclusion and exclusion criteria. A maximum of 55 UK NHS Trusts/Health Boards are involved in trial recruitment, enrolling 740 patients who will be randomly allocated to receive either carvedilol or a matched placebo in a 1:1 ratio and treated for 36 months (or up to a minimum of 185 events), with a follow-up visit every 6 months during this period. Participants will also be followed up until last patient last visit, by record linkage to the Hospital Episodes Statistics (HES) electronic dataset in England and from participating hospital records in Wales, Northern Ireland, and Scotland. Death records held by the Office for National Statistics will also be used to assess the impact of the trial medication period on long-term outcomes.

**Figure 1.**
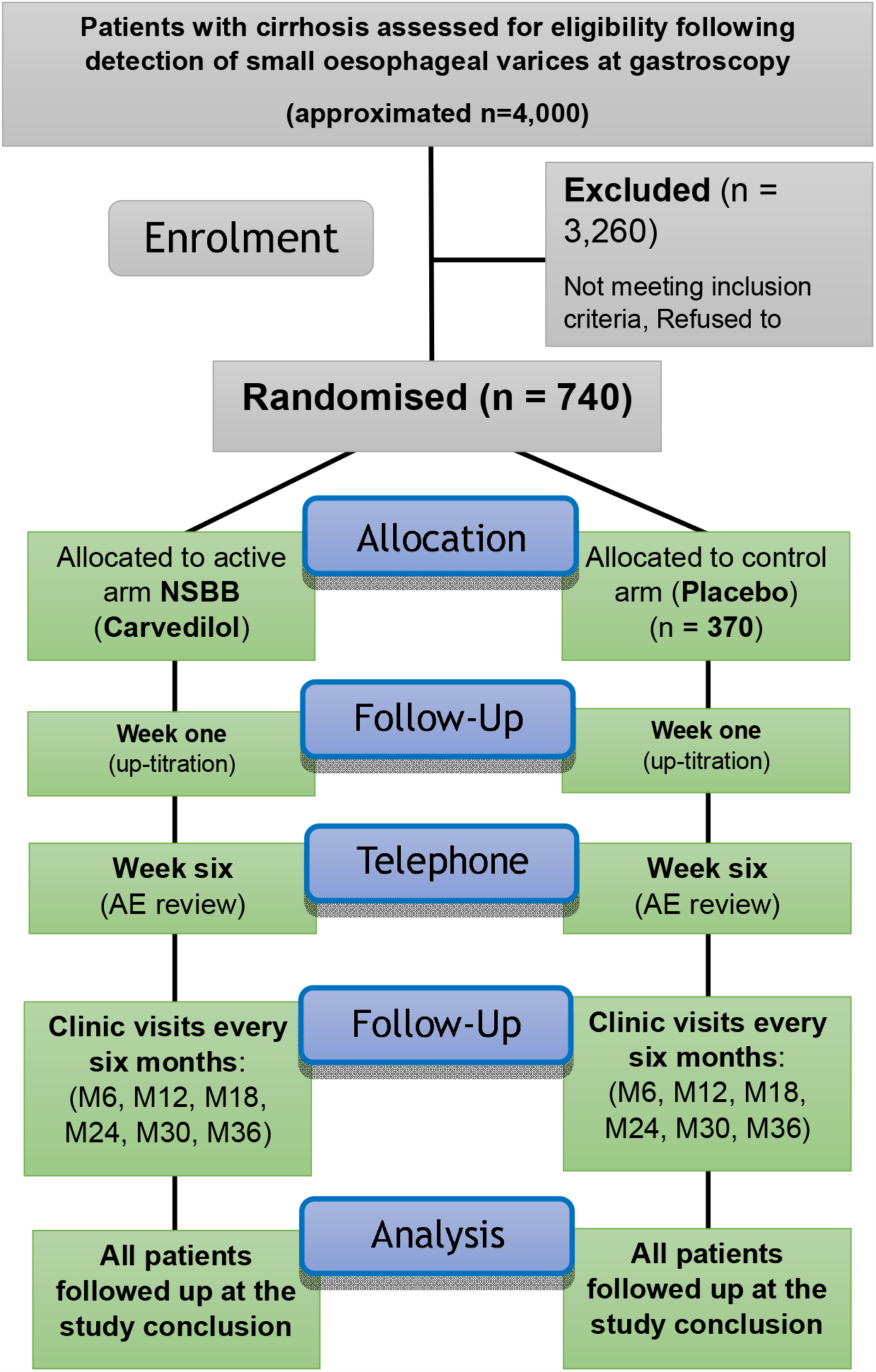
Trial schema

### Follow-up completion

Last patient last visit (LPLV) will take place after a minimum of 36 months follow-up or may be triggered by DMC recommendation and ratified by TSC when a minimum of 185 events is reached.

## Methods

### Study setting

BOPPP will take place at NHS hospitals in the UK, who manage patients with cirrhosis. A list of study sites can be found in the Appendix 2. Patients with cirrhosis and small oesophageal varices that are detected whilst attending variceal screening gastroscopies, will be identified, pre-screened, approached to participate.

### Eligibility criteria

To be eligible for BOPPP, a patient must have cirrhosis and small oesophageal varices (as a manifestation of portal hypertension) detected by endoscopy within 6 months of being recruited. Small or grade I oesophageal varices are characterised as veins in the oesophagus which are ≤5 mm in diameter and/or veins that completely disappear upon moderate air insufflation at gastroscopy. The complete eligibility criteria for the trial is outlined in Table 2.

**Table 2.** Eligibility criteria

|  |
| --- |
| **Inclusion criteria**<br>**Cirrhosis and portal hypertension, defined by any 2 of the following:**<br>- Characteristic clinical examination findings; one or more of<br>- characteristic liver function tests<br>- haematological panel<br>- coagulation profile abnormalities<br>- Characteristic radiological findings; one or more of<br>- heterogeneous liver with irregular contour<br>- Splenomegaly<br>- ascites<br>- varices<br>- recanalized umbilical vein<br>- FibroScan liver stiffness measurement >15 kPa without other explanation<br>- Fibrosis score > ISHAK stage 4 on liver biopsy<br>**Small oesophageal varices diagnosed within the last 6 months\***<br>**Not received a beta-blocker in the last week**<br>**Capacity to provide informed consent** |
| **Exclusion criteria**<br>**Non-cirrhotic portal hypertension**<br>**Current medium/large oesophageal varices (defined as >5 mm in diameter)**<br>**Previous medium/large oesophageal varices, which decreased in size with curative therapy**<br>**Gastric (IGV and GOV2), duodenal, rectal or other ectopic varices with or without evidence of recent bleeding. For gastric varices, this includes: IGV-1 and IGV-2 (isolated gastric varices) and GOV2 (gastric varices continuing into the cardia).**<br>• *[GOV1 (gastric varices continuing into the lesser curve) are not an exclusion if present with small oesophageal varices]*<br>**Previous variceal haemorrhage**<br>**Previous band ligation or glue injection of oesophageal and/or gastric varices**<br>**Red signs accompanying small oesophageal varices at endoscopy**<br>**Known intolerance to beta blockers**<br>**Contraindications to beta blocker use:** Heart rate <50 bpm, known 2nd degree or higher heart block, sick sinus syndrome, systolic blood pressure <85 mmHg, chronic airways obstruction (asthma/COPD), Floppy Iris Syndrome, CYP2D6 poor metaboliser, history of cardiogenic shock, history of severe hypersensitivity reaction to beta-blockers, untreated pheochromocytoma, severe peripheral vascular disease, Prinzmetal angina and NYHA IV heart failure<br>**Unable to provide informed consent\***<br>**Child Pugh C cirrhosis\***<br>**Already receiving a beta-blocker for another reason that cannot be discontinued**<br>**Graft cirrhosis post liver transplantation**<br>**Evidence of active malignancy without curative therapy planned**<br>**Pregnant or lactating women\***<br>**Women of child bearing potential not willing to use adequate contraception during the period of IMP dosing (if relevant)**<br>**Patients who have been on a CTIMP within the previous 3 months**<br>**Clinical symptoms consistent with COVID-19 at the time of randomisation** |

\*Due to the dynamic nature of these measures in such investigations, patient's condition with cirrhosis may change. Therefore, patients may be re-assessed for eligibility.

Upon analysis of pre-screening data from participating sites, a significant proportion of potentially eligible patients were being excluded from the trial in having a diagnosis of asthma or chronic obstructive pulmonary disease (COPD) documented on their medical history. To mitigate this, the exclusion criterion “contraindication to beta-blocker use due to asthma/COPD” was clarified in the protocol to exclude only patients with a potential risk of side effects.

### Recruitment and informed consent

A participant information sheet (PIS) is given to and discussed with potential patient recruits before informed consent is sought. Written informed consent is obtained by medical staff who are delegated at each site and have received protocol-specific training. Only patients aged 18 and above are considered for consent.

### Randomisation and participant timeline

The electronic data capture database, InferMed MACRO is used to register patients. The system generates a unique trial patient identification number (PIN). Randomisation is provided by a secure 24 hours web-based randomisation service. The King’s Clinical Trials Unit (KCTU) hosts both MACRO and the randomisation service.

The allocation sequence was stratified block randomisation with randomly varying block sizes. The KCTU IMP pharmacy management system assigns the patient to a treatment group and generates anonymised treatment pack numbers for trial staff to identify the correct trial medication for the patient, whilst maintaining blinding.

Participants who have been randomised to either treatment, begin with one tablet a day of 6.25mg carvedilol or placebo. Patients are expected to start the trial medication on the same day as randomisation or as soon as possible thereafter. A face-to-face follow-up visit is scheduled for haemodynamic review at week 1 with a view of up-titration to 2 tablets (12.5mg carvedilol or placebo) if participants meet the outlined parameters in Appendix 1. Subsequently, a safety telephone call is completed at week 6. Thereafter, participants are seen in during outpatient appointments at 6-monthly intervals over the 3-year follow-up period. Month 36 marks the end of the trial treatment period and denotes the minimum trial follow-up period. Patients who complete the treatment period prior to last patient last visit (LPLV) will be followed up once again at the end of the trial.

### Schedule of events

This is detailed in Table 3.

**Table 3.** Schedule of events

| Trial procedures | Pre-screening | Screening Visit | Baseline | Week 1 (+/-) 3 days | Week 6 (+/-) 2 weeks | Month 6 (+/-) 6 weeks | Month 12 (+/-) 6 weeks | Month 18 (+/-) 6 weeks | Month 24 (+/-) 6 weeks | Month 30 (+/-) 6 weeks | Month 36 (+/-) 6 weeks | At variceal bleed | At trial completion (via notes) |
| --- | --- | --- | --- | --- | --- | --- | --- | --- | --- | --- | --- | --- | --- |
| Informed consent |  | X |  |  |  |  |  |  |  |  |  |  |  |
| Eligibility criteria |  | X | X |  |  |  |  |  |  |  |  |  |  |
| Randomisation |  |  | X |  |  |  |  |  |  |  |  |  |  |
| Demographics* |  | X |  |  |  |  |  |  |  |  |  |  |  |
| Medical History* |  | X |  |  |  |  |  |  |  |  |  |  |  |
| Targeted physical exam* |  | X | X*** |  |  | X | X | X | X | X | X |  |  |
| Weight / Height* |  | X | X*** |  |  | X | X | X | X | X | X |  |  |
| Vital signs (BP/HR)* |  | X | X | X |  | X | X | X | X | X | X | X |  |
| TE/APRI (FibroScan)* | X |  |  |  |  |  |  |  |  |  |  |  |  |
| FBC, INR, Liver, Renal and Bone profile* |  |  | X† |  |  | X | X | X | X | X | X | X |  |
| Liver prognostic scores* L |  |  | X† |  |  | X | X | X | X | X | X | X |  |
| AUDIT-C/alcohol questionnaire* |  |  | X |  |  | X | X | X | X | X | X |  |  |
| Variceal Haemorrhage Status |  |  |  |  |  | X | X | X | X | X | X | X |  |
| IMP dispensing |  |  | X<sup>a</sup> |  |  | X | X | X | X | X |  |  |  |
| Commence IMP |  |  | X |  |  |  |  |  |  |  |  |  |  |
| Dose - titration |  |  |  | X | X | X | X | X | X | X | X | X |  |
| HCC surveillance US* | X<sup>‡‡</sup> |  |  |  |  | X | X | X | X | X | X | (X) |  |
| Gastroscopy* | X† |  |  |  |  |  | X |  | X |  | X | X |  |
| Conmeds* |  |  | X | X | X | X | X | X | X | X | X |  |  |
| Adverse events (AEs)** |  |  |  | X | X | X | X | X | X | X | X |  | X |
| QoL questionnaire |  |  | X |  |  | X | X | X | X | X | X |  |  |
| Health Care Usage |  |  | X |  |  | X | X | X | X | X | X |  | X |
| Adherence to IMP |  |  |  | X |  | X | X | X | X | X | X | X |  |
| Telephone call |  |  |  |  | X |  |  |  |  |  |  |  |  |

**Note** \*standard of care ; \*\*AEs collected from baseline to 30-days post M36 visit/ permanent IMP discontinuation (not including death), \*\*\*not repeated if screening and baseline are within 2 weeks, ‡‡ completed within 6 months of screening or last SOC surveillance for HCC-US [alternative imaging methods [CT and / or MRI] are permitted as long as the data needed is provided]), † completed within 6 months of baseline, □ Child-Pugh, MELD, UKELD, CLIF-C AD, α IMP to be allocated within 4 weeks of consent

### Trial intervention, dosing regimen and dose modification

The Investigational Medicinal Products (IMPs) are carvedilol 6.25mg and a matched placebo tablet. The IMP is taken once daily from randomisation and can be up-titrated to a maximum daily dose of 12.5 mg (2 tablets once daily or 1 tablet twice daily) following a haemodynamic review at week 1. The criteria for dose modification is listed in Appendix 1. Patients are instructed to take one or two tablets per day for a total period of 3 years.

Carvedilol is generally well-tolerated resulting in a fewer compliance issues or lower rate of discontinuation due to adverse events (AEs) than other NSBBs (6). Guidance for temporary and permanent cessation of IMP is detailed in Figure 2. Where required, the dose will be up- or down-titrated at clinician discretion, at trial visits and if the patient contacts the trial team regarding side effects.

**Figure 2.**
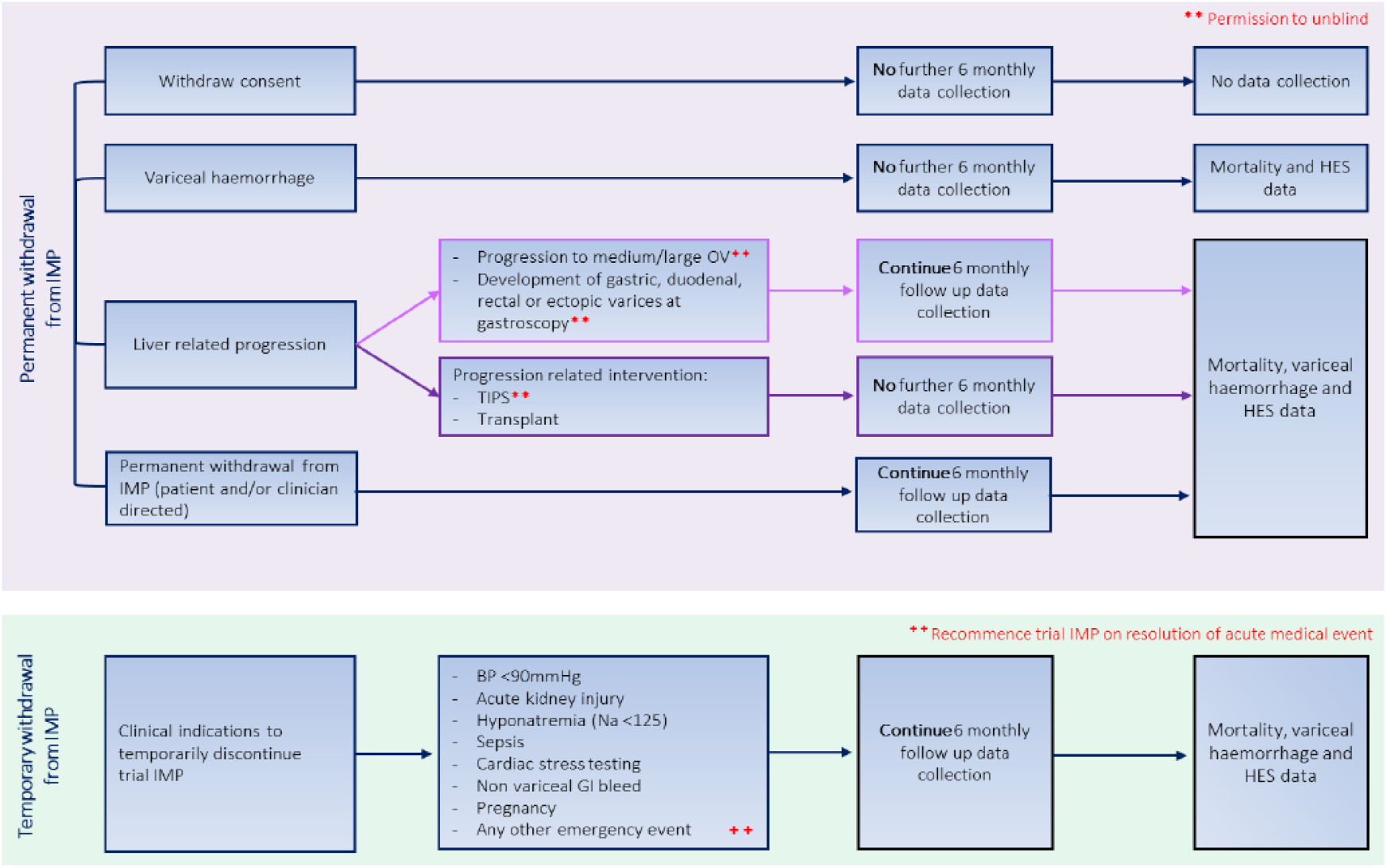
Pathway of follow-up procedures or events resulting in permanent or temporary discontinuation of the trial Investigational Medicinal Product (IMP).

A safety telephone call is arranged at week 6 to assess short-term adverse events such as hypotension, gastrointestinal side effects like nausea, swelling of hands and feet, blurred vision, lethargy, headache, sexual dysfunction and shortness of breath. Participants are asked about their adherence to the trial medication at each follow-up.

### Blinding

#### Who will be blinded?

Trial participants, trial staff (including clinicians and on-site pharmacy teams) and the trial analyst will be blinded. The carvedilol and placebo tablets are identical in appearance. A junior trial statistician will be planned to be partially blinded (able to access outcome data labelled as A or B) during the study to provide data split by arm to the Data Monitoring Committee (DMC).

#### Procedure for unblinding

There will be no unblinding unless deemed emergent for the patient’s care as assessed by the attending clinicians. In the event of emergency unblinding, a 24-hour telephone unblinding service is provided for Emergency Code Break and Medical Information by ESMS Global Ltd. All randomised participants will be provided with an alert card detailing a code break telephone number and emergency contact details. All randomised participants are requested to carry the alert card with them at all times whilst participating in the trial.

### Data collection and management

The data will be collected using source data worksheets and will be transcribed into an electronic data capture database (InferMed MACRO). The follow-up schedule is pragmatically designed to align with regular clinical follow-up appointments to minimise the additional burden for trial participants, thus promoting retention.

Pre-screening, screening and enrolment logs will be kept at each trial site in a secured room either locked in a cabinet or electronically. Electronic data will be stored on secure servers based at the lead organisation. The databases will be password-protected and only accessible to specified and delegated trial individuals. After completion of the trial, these logs will be archived and stored securely in an archiving facility for a minimum of 15 years.

### Primary outcome

- Time to first decompensating event as defined by Table 1
- Cost effectiveness of carvedilol in this population.

*For the purpose of inferential statistical inference, only all-cause decompensation will be defined as the primary outcome, analysed with a two sides hypothesis test (α = 0*.*05)*

### Secondary and tertiary outcomes

- Estimation of the 1, and 3-year oesophageal variceal bleed rate by allocation, and associated number needed to treat
- Progression to oesophageal medium/large varices requiring clinical intervention over 3 years
- Composite of oesophageal variceal bleed or progression to medium/large varices over 3 years
- Development of gastric, duodenal, ectopic or rectal varices
- Survival (Overall, liver-related, or cardiovascular-related)
- Quality of life, using the EQ-5D-5L questionnaire

#### Qualitative interviews to understand recruitment barriers and enablers

During the first 12 months, semi-structured interviews were conducted which examined views and experiences of the proposed intervention, trial recruitment and trial procedures with patients who enrolled in the trial (n=12), patients who declined to take part (n=5), and staff who were responsible for recruiting participants to the trial (n=18). Potential barriers and facilitators to recruitment were considered at the level of the patient, staff, team, organisational and trial (across organisations). Recruitment was conducted across 14 NHS Trusts, chosen to provide a mix of regions and recruitment success, and continued until thematic saturation was reached. (See Le Boutillier et al., 2022a for a full description of the procedures(16)).

#### Qualitative interviews to understand barriers and enablers to understand future implementation in primary care

In the first 12 months of BOPPP, attention was given to early engagement of GPs in conversations around the delivery of this potentially effective secondary care-initiated treatment in primary care to support future implementation. Semi-structured individual interviews explored GP perspectives (n=23) on factors that could influence implementation beyond the trial and how dose titration and ongoing treatment with carvedilol could be best delivered in primary care. GPs were recruited through ten Clinical Commissioning Groups (CGGs), selected purposively to include a mix of regions and practice size.(17)

### MBOP – Mechanism of beta-blockade on bacterial translocation in portal hypertension (MBOP) sub-study

An integrated basic science mechanistic study has been established to investigate the mechanism of carvedilol in preventing ACD in patients with cirrhosis. All BOPPP sites are offered the opportunity to participate in the MBOP sub-study, where BOPPP participants, at baseline, are separately consented to provide biological samples longitudinally. This work will be reported outside of the primary results manuscript.

### Statistical methods

A statistical analysis plan (SAP) was drafted by the trial statistician (HJ) and senior statistician (BC) who were blinded to outcome data at time of drafting. The SAP was approved by the TSC chair and an independent statistician. [Supplementary document 1]

### Sample size estimation

The PREDESCI study enrolled 201 patients (1:1) and reported a 3-year decompensation rate of 27% in the placebo group, compared to 16% in the β-blockers group (hazard ratio [HR] 0.51, 95% confidence interval [CI] 0.26–0.97, p=0.04).

The BOPPP trial, with 187 patients randomized as of December 2021, exhibited an annual all-cause decompensation event rate of approximately 0.16 across both treatment arms. Additionally, another data extract from the BOPPP database in March 2022 showed that the 1-year all-cause decompensation rate was 20% across both arms combined. Anticipating a conservative escalation in event rate over a three-year span, and to accommodate attrition due to censoring, due to adverse events causing treatment discontinuation in these patients, we project a minimum decompensation rate of 25.5% for the trial duration. This estimate shows a projected rate of 31% in the placebo arm and 20% in the β-blocker arm. For detecting the postulated difference, which corresponds to a hazard ratio of 0.60, with a statistical power of 90% and a type I error probability of 0.05, a sample size of 666 patients resulting in 170 events is required. To anticipate a dropout rate of 10%, an enrolment target of 740 patients is set.

### Sample size justification and power analysis

In estimating the required sample size for our study, we based calculations on an anticipated hazard ratio (HR) of 0.6. However, should the true HR prove to be 0.7, with the control arm event rate remaining constant, our power analysis indicates that the trial would still maintain a power of 71%. This is based on a conservative event rate estimate of 25.5% for both arms. We have reason to believe that the actual event rate may exceed this conservative projection. Consequently, we assert that the study is adequately powered to detect a clinically significant effect, even if the true hazard ratio is somewhat higher than initially anticipated.

### Internal pilot

At 12 months after participant recruitment opens, we assessed the feasibility of recruitment and retention acceptability, with progression criteria: 1) at least 8 sites opened, with at least one patient randomised at each. 2) at least 80 patients randomised, and 3) at least 70% retention rate. Upon review at the 12-month milestone, all progression criteria were fulfilled—evidencing effective site activation, patient randomization, and retention rates—and the results have been duly reported to the Data Monitoring Committee (DMC) and Trial Steering Committee (TSC).

### Population under investigation

The intention-to-treat (ITT) population will include all patients enrolled in the study and involved in the assessment of outcomes. Patients experiencing any of all-cause decompensation events, as listed in Table 1, whichever occurs first, will be classified as treatment failures, and recorded as events. Those who do not experience any form of decompensation during the follow-up period will be considered non-events and censored at their last follow-up time.

The per-protocol population (PPP) comprises participants who adhere to the study protocol.

### Protocol deviations and violations

A protocol deviation or violation refers to any unplanned divergence from the planned study protocol. A protocol deviation (PD) is characterized as a minor, non-serious departure from the protocol that is unlikely to affect the integrity of the data or the overall treatment effect. An instance of a PD may include the missing of a scheduled visit within the allowable window or the failure to return the IMP bottle during a visit.

Conversely, a protocol violation (PV) represents a more significant departure from the protocol that has the potential to substantially impact the quality or interpretability of the data. Such violations may result in the exclusion of the affected patient from the per-protocol population. Comprehensive definitions and categorizations of protocol deviations and violations have been elaborated upon in the SAP.

### Adherence to trial intervention

Adherence will be defined in the SAP.

### Recruitment, descriptives and baseline comparability

In this study, the flow of participants from screening through to 36-month follow-up will be presented in a CONSORT flowchart, ensuring transparency and accountability of reporting clinical trial data. This includes the number of individuals screened, reasons for ineligibility, and retention rates throughout the follow-up period. Descriptive statistics will be used to summarise evaluations of recruitment, drop-out, and therapy completion, as well as demographics and primary and secondary outcomes comparability at baseline.

### Analysis of the primary outcome

The primary analysis will focus on the time to all-cause decompensation between the two allocated groups. To assess the impact of treatment on decompensation risk over time, we will employ a multi-level Cox proportional hazards regression model. This model will adjust for covariates including patient age, gender, and MELD score, which are recognized as potential confounders. To account for hospital variability, we will incorporate site as a shared frailty across hospitals within the model. The proportional hazards assumption will be examined through visual inspection of Kaplan-Meier plots and log-log survival plots, complemented by an at-risk table for both treatment groups. The primary measures of effect will be the adjusted hazard ratios (aHR), accompanied by 95% confidence intervals and p-values. Analysis will be carried our using a two sided test (*α* = 0.05).

All analyses for secondary endpoints will be conducted within the intention-to-treat (ITT) population. No adjustment will be made for multiple secondary outcomes (*α* = 0.05). Binary outcomes such as variceal size progression (progressed versus not progressed), will be evaluated through generalized linear mixed-effects logistic regression models at both 1-year and 3-year post-randomization intervals. In this model site will be incorporated as a random effect. Adjustments will be made for covariates including age, sex, aetiology of liver disease (alcoholic, non-alcoholic fatty liver disease (NAFLD), viral hepatitis, autoimmune, or other), and disease severity as measured by the MELD score at baseline.

Time-to-event secondary outcomes will be defined through hazard ratios with 95% confidence intervals and visualized via Kaplan-Meier survival plots. Median times to event occurrence and their two-sided 95% confidence intervals will also be reported. Cox proportional hazards models similar to primary outcome will be used for estimation of aHR and their 95% confidence intervals.

Continuous variables, such as deterioration in liver function, will be measured using the Child-Pugh and MELD scores. A Linear Mixed Model (LMM) will compare these outcomes between treatment groups, accounting for site variability as a random effect. Changes over time in these scores will be analysed by incorporating visit time-points (baseline and every six months up to 36 months) as time-varying covariates. Subject ID will be included to account for intra-participant correlation, in addition to site as a random effect. The model will be adjusted for baseline covariates. Secondary outcomes may not be powered to detect statistical differences.

### Sensitivity analyses

Sensitivity analyses will be performed to determine the robustness of the trial’s conclusions to the potential effects of missing data and non-adherence. These analyses will be conducted to provide supplementary exploratory assessments for both primary and secondary endpoints

### Cost effectiveness analysis (economic evaluation)

A separate Health Economics Analysis Plan (HEAP) will be produced, providing a comprehensive description of the planned economic evaluation. Briefly, the primary objective of the health economic evaluation is to calculate the cost-effectiveness of carvedilol versus placebo control over 3 years in a within-trial economic evaluation. Secondary objective is to calculate the cost-effectiveness of carvedilol versus placebo over a lifetime using economic modelling techniques. Data on community healthcare services used by the participants to estimate costs will be collected every 6 months including baseline using self-completed questionnaire and covering the 3 years period. Hospital-based service use will be obtained from the Hospital Episode Statistics records in England or from participating hospitals in Scotland, Wales, or Northern Ireland, and intervention related resource use (medication) will be obtained from IMP adherence log.

Data on health-related quality of life, using measures capable of generating quality adjusted life years (QALYs) for use in economic evaluations, will be collected using the EQ-5D-5L measure(18). The EQ-5D-5L measure consists of 5 questions (covering mobility, self-care, usual activities, pain/discomfort and anxiety/depression), each with 5-level responses. The measure will be completed at baseline, 6-, 12-, 18-, 24-, 30-, 36-months post randomisation. Utility values for each health state at each time point will be estimated by means of a mapping to the available EQ-5D-3L set of preference weights using population values. QALYs will be estimated(19) for the defined period using a linear interpolation to calculate the area under the QALY curve.

In line with the clinical analyses, analysis will be ‘as randomised’ (intention-to-treat), where participants are analysed according to their allocation, regardless of whether they received that treatment or not. The primary economic analysis will take the health and personal social services (PSS) perspective and will explore the cost-effectiveness of carvedilol compare with placebo at 36-months in terms of cost per QALYs. A secondary economic analysis will extend this to compare costs and QALYs modelled for a lifetime after the end of the RCT. The mean difference in total cost and QALYs per participant between the randomised arms will be estimated using bootstrapped regression, adjusted in line with the clinical analyses, plus the baseline variable of interest (baseline cost and/or baseline utility score).

We will examine cost-effectiveness through incremental cost-effectiveness ratios (ICERs) for any combinations of cost and outcome that involve a trade-off, where one group incurs both higher costs and greater benefits compared to the other. Combinations with lower costs and higher outcomes are considered ‘dominant.’ The cost-effectiveness of an intervention is determined by whether the ICER value is above or below the National Institute of Health and Care Excellence (NICE) willingness to pay per QALY threshold of £20-£30,000. Additionally, we will represent cost-effectiveness using incremental net health benefit (INB). The INB indicates the adjusted mean difference in benefit in terms of QALYs score by transforming the adjusted mean difference in total cost between the intervention and the control onto the QALYs scale using the specified threshold value. In contrast to ICER, where interpretation depends on the direction of the incremental cost and effect, the interpretation of INB is straightforward: the intervention is deemed cost-effective if its INB is positive.

Uncertainty around the cost-effectiveness analysis will be explored using cost-effectiveness planes and cost-effectiveness acceptability curves (CEACs)(20). Cost-effectiveness planes plot the mean differences in total cost and QALYs. The cost-effectiveness acceptability curve will be derived by calculating the proportion of bootstrapped estimates that are cost-effective across a range of willingness-to-pay thresholds, to show the probability that the intervention is cost-effective across different threshold values.

### Analysis of qualitative data

Qualitative data was transcribed verbatim. Inductive thematic analysis was used to identify themes of importance to patients and to staff who were responsible for recruiting patients to the trial (21). Separate coding frames were initially developed for patients and staff through a process of line-by-line coding, organising, and reviewing themes. Interpretative analysis was then undertaken to identify overarching themes, which were mapped to the Theoretical Domains Framework (TDF) to provide a theoretical understanding of factors that influence recruitment practices (22). Based on this framework, we identified 16 strategies that could be used to support recruitment by addressing the barriers and enhancing the enablers to recruitment, which were then discussed, prioritised and implemented among the research team(16).

Data from GP interviews were analysed using reflexive thematic analysis (23), ensuring a thorough engagement with the data and depth of interpretation. Following a process of data familiarisation, codes were identified through line-by-line coding, refined, and grouped into preliminary themes on the basis of shared ideas or concepts. Refinements to the specifics of themes, and thematic patterns continued until a useful and meaningful analysis was achieved.

### Missing data and sensitivity analyses

Every attempt will be made to collect full follow-up data on all study participants, and it thus anticipated that missing data will be minimal. Patients who withdraw from IMP will be invited to continue follow up where possible.

#### Confidentiality

All data will be handled in accordance with the Data Protection Act 2018.

## Trial organisation, ethics and dissemination

### Trial Management Group

The Trial Management Group (TMG) is responsible for assisting with the design, coordination and day-to-day operational and strategic management of the trial. The TMG is composed of Chief Investigator (CI), Chief Scientific Investigator (CSI), expert clinicians (hepatologists, gastroenterologists), trial methodologist, statistician, trial manager, trial pharmacist, qualitative researchers, CRN representatives, patient advocates and health economists.

### Data Monitoring Committee

The Data Monitoring Committee (DMC) is comprised of an independent clinical chair with expertise in liver disease, an independent hepatologist, and an independent statistician. The DMC will have access accumulating comparative data and in accordance with ICH-GCP guidelines, are responsible for monitoring trial conduct and safety, assessing risk and benefits and making recommendations to safeguard the interests of trial participants. The DMC will inform the Trial Steering Committee (TSC) if there are any issues raised from its discussions and make recommendations on trial continuation based on adverse events and adverse reactions reported.

### Trial Steering Committee

The Trial Steering Committee (TSC) will assess trial conduct and recruitment. The TSC is comprised of an independent clinical chair, chief investigator, trial statistician, clinical co-applicants, patient representatives and independent primary and secondary care clinicians. This group oversee the running of the trial and discuss any issues that may arise throughout recruitment and follow-up of participants. If the DMC make a recommendation to prematurely terminate the trial, the TSC will review and decide course of action

### Ethical approval

Ethical approval was provided by the Leeds West Research Ethics Committee, (Reference: 19/YH/0015) and a clinical trials authorisation issued by the Medicines and Healthcare products Regulatory Agency (MHRA), (Reference: CTA 21416/0243/001-0001)

### Frequency and plan for monitoring trial conduct

To ensure patient safety and data integrity, all sites will be monitored (on-site or remote) to ensure quality assurance in that the site is adhering to the trial protocol, Good Clinical Practice (GCP) guidelines and regulations, during the running of the trial. The first monitoring visit, following initiation of the site and trial commencement will take place within 8 weeks of randomising the first patient. Subsequent monitoring visits will take place every 6 months thereafter. In addition, the sites will be centrally monitored were study data will be regularly checked for any anomalies.

### Dissemination

Results of this trial will be submitted for publication in a peer-reviewed scientific journal to maximise chances of acceptance and implementation into clinical practice, regardless of effect on outcomes. The manuscript will be prepared by CI or delegate and authorship will be determined by the trial publication policy. Research findings will also be presented at conferences and seminars. Participants will be informed of the results via the BOPPP website (www.boppp-trial.org) and partners in the British Liver Trust (BLT) organisation.

## Trial status

The BOPPP trial began recruitment in August 2019 and recruitment is ongoing. Due to the COVID-19 pandemic, recruitment was paused, which affected trial timelines. The anticipated recruitment end date is now mid-2024.

## Supporting information

SAP

## Data Availability

The main outcomes will be published in a peer-reviewed journal. Unidentifiable participant data will be presented in the trial results. The full study protocol and statistical analysis plans can be requested by email to

## Abbreviations

BOPPP: Beta-blockers or Placebo for Primary Prophylaxis of oesophageal varices
MBOP: Mechanism of beta-blockade on bacterial translocation in portal hypertension
GCP: Good Clinical Practice
CI: Chief Investigator
CSI: Chief Scientific Investigator
CRN: Clinical Research Network
TMG: Trial Management Group
DMC: Data Monitoring Committee
TSC: Trial Steering Committee
PPI: Personal and Public Involvement
BLT: British Liver Trust
NSBB: Non-selective beta-blocker
ACD: All-cause decompensation
RCT: Randomised controlled trial
MELD: Model for End-Stage Liver Disease
OV: Oesophageal varices
VH: Variceal Haemorrhage
NHS: National Health Service
REC: Research Ethics Committee
MHRA: Medicines and Healthcare products Regulatory Agency
CSPH: Clinically significant portal hypertension
NICE: National Institute of Health and Care Excellence
BSG: The British Society of Gastroenterology
COVID-19: Coronavirus disease
PREDESCI: β blockers to prevent decompensation of cirrhosis in patients with clinically significant portal hypertension
SAP: Statistical Analysis Plan
HEAP: Health Economics Analysis Plan
NIHR: National Institute for Health and Care Research
HTA: Health Technology Assessment
LPLV: Last patient last visit
UK: United Kingdom
IGV: Isolated gastric varices
GOV: Gastroesophageal varices
CTIMP: Clinical Trial of an Investigational Medicinal Product
COPD: Chronic obstructive pulmonary disease
PIS: Patient Information Sheet
KCTU: King’s Clinical Trials Unit
IMP: Investigational Medicinal Products
GP: General Practitioner
aHR: adjusted hazard ratios
ITT: intention-to-treat
PPP: per-protocol population
CONSORT: Consolidated Standards of Reporting Trials
NAFLD: Non-alcoholic fatty liver disease
LMM: Linear Mixed Model
QALY: quality adjusted life years
ICER: incremental cost-effectiveness ratios
INB: incremental net health benefit
CEAC: cost-effectiveness acceptability curves
TDF: Theoretical Domains Framework

## Declarations

### Consent for publication

Not applicable – no identifying images or personal or clinical details of participants are presented here or will be presented in reports of the trial results.

## Acknowledgements

The BOPPP trial team are grateful to all trial participants, without whom this trial would not be possible. We also acknowledge the contributions of the following:

- Independent members of the Trial Steering Committee: Prof Eleanor Barnes, Dr Sam Thomson, Dr Charles Millson, Dr Oliver Van Hecke, Prof Alan Watkins, Dr Claire Snowdon, Elaine Mullings, Morwenna Orton and Peter Walsh. *We would like to thank a former member of the TSC, Neville Hargrave, who contributed hugely to the design and conduct of the trial but unfortunately passed away in 2022*.
- Independent members of the Data Monitoring Committee: Kenneth Simpson, Jeremy Cobbold, Stephanie MacNeill
- NIHR local clinical research networks
- All PIs and trial staff at all recruiting sites (listed under BOPPP study group [Supplementary document 2])

## Authors’ contributions

VP is the chief investigator and MM is the chief scientific investigator; both conceived the study and led the proposal. BC is the lead statistician and methodologist; he designed the study and is leading the analysis. RU manages the project. HJ developed the statistical analysis plan alongside BC and reviewed the study design. VL and CLB designed the qualitative element of the trial and the corresponding section of the manuscript. NY and JS contributed to the design of the health economic analysis of the trial and the corresponding section of the manuscript. VP, MM, BC, HJ, and RU contributed to the development of the protocol. All authors read, edited, and approved the final manuscript.

## Trial funding and disclaimer

The trial was commissioned and is funded by the UK National Institute for Health and Care Research Health Technology Assessment Board (NIHR-HTA): Award ID: 17/32/04 https://www.fundingawards.nihr.ac.uk/award/17/32/04; the funding body was not involved in the data collection and processing, or the writing of this manuscript. The views expressed are those of the author(s) and not necessarily those of the National Health Service (NHS), the NIHR or the Department of Health and Social Care

## Appendix 1

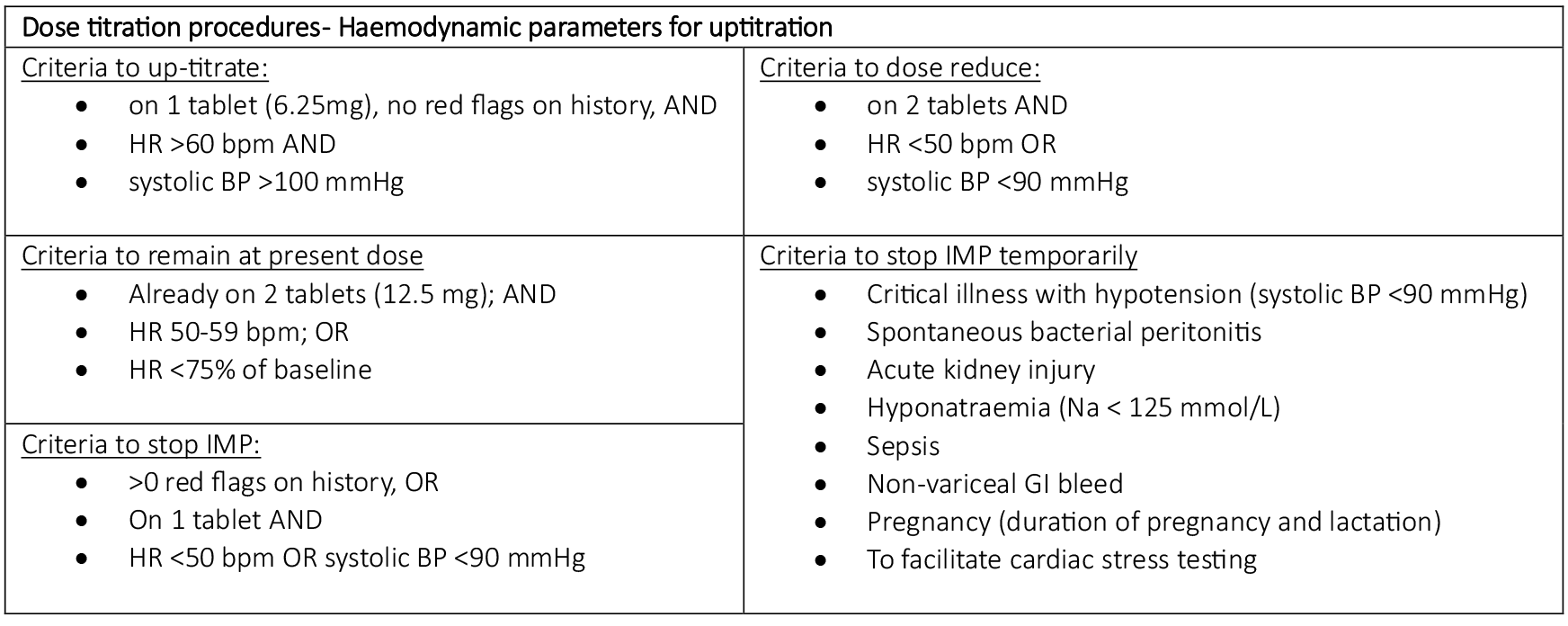

| Dose titration procedures- Haemodynamic parameters for uptitration |  |
| --- | --- |
| <u>Criteria to up-titrate:</u><br>• on 1 tablet (6.25mg), no red flags on history, AND<br>• HR >60 bpm AND<br>• systolic BP >100 mmHg | <u>Criteria to dose reduce:</u><br>• on 2 tablets AND<br>• HR <50 bpm OR<br>• systolic BP <90 mmHg |
| <u>Criteria to remain at present dose</u><br>• Already on 2 tablets (12.5 mg); AND<br>• HR 50-59 bpm; OR<br>• HR <75% of baseline | <u>Criteria to stop IMP temporarily</u><br>• Critical illness with hypotension (systolic BP <90 mmHg)<br>• Spontaneous bacterial peritonitis<br>• Acute kidney injury<br>• Hyponatraemia (Na < 125 mmol/L)<br>• Sepsis<br>• Non-variceal GI bleed<br>• Pregnancy (duration of pregnancy and lactation)<br>• To facilitate cardiac stress testing |
| <u>Criteria to stop IMP:</u><br>• >0 red flags on history, OR<br>• On 1 tablet AND<br>• HR <50 bpm OR systolic BP <90 mmHg |  |

## Appendix 2

BOPPP study group. List of sites and collaborating authors.

1. Aberdeen Royal Infirmary - Dr Ashis Mukhopadhya, Alicija Vileito, Tracy Henderson
2. Addenbrooke’s Hospital - Dr Gwilym Webb, Jerrian Joyce Andrada, Abigail Ford
3. Aintree University Hospital - Dr Cyril Sieberhagen, Claire Burston, Carol Brooks
4. Basildon University Hospital - Dr Gavin Wright, Bushena Miyesa, Aimee Williams
5. Bedford Hospital - Dr Jay Patel, Melchizedek Penacerrada
6. Bristol Royal Infirmary - Dr Gautham Appanna, Gifthy Perez, Joanne Elliott
7. Broomfield Hospital - Dr Keval Naik, Susan Smolen, Anna Beckwith
8. Chelsea and Westminster Hospital - Dr Matthew Foxton, Carina Bautista
9. Derriford Hospital - Prof Matthew Cramp, Ada Laureen Nweze
10. East Surrey Hospital - Dr Gayatri Chakrabarty, Indhuja Rajkumar, Merlin James
11. Freeman Hospital - Dr Steven Masson, Sheenu Thomas, Lucy Dixon, Sarah Hogg, Louise Finlay
12. Frimley Park Hospital - Dr Kuldeep Cheent, Jessica Camp
13. Glasgow Royal Infirmary - Prof Adrian Stanley, Alexis Duncan, Lauren Walker
14. Gloucestershire Royal Hospital - Dr Duncan Napier, Paula Hilltout, Linda Hill, Hiromi Uzu
15. Great Western Hospital - Dr Moby Joseph, Suzannah Pegler, Camille Walling
16. Hull Royal Infirmary - Dr Lynsey Corless, Anisoara Kingsbury, Tania Nurun
17. Kettering General Hospital - Dr Debasish Das, Anna Williams
18. King’s College Hospital - Dr Mark McPhail, Ane Zamalloa
19. King’s Mill Hospital - Dr Stephen Foley, Camelia Goodwin
20. Kingston Hospital - Dr Markus Gess, Margaret Grout
21. Leicester General Hospital - Dr Ka-Kit Li, Olivia Watchorn, Laura Plummer
22. Lewisham General Hospital - Dr Laura Blackmore, Christos Tsintikidis, Allysha Perryman
23. Maidstone Hospital - Dr George Bird, Emily Phiri
24. Medway Maritime Hospital - Dr Mohamed Saleh, Dr Adaze Woghiren, Dilukshi Wickramasinghe, Jodie Wright
25. Ninewells Hospital (Tayside) - Dr Michael Miller, Shona Murray, Leanne Cosgrove
26. Pinderfields Hospital - Dr John Hutchinson, Julie Burton, Emma Stoner, Stephanie Lupton
27. Princess Royal University Hospital - Dr Mayur Kumar, Nicola Griffiths, Anna Posada
28. Queen Alexandra Hospital (Portsmouth) - Dr Andrew Fowell, Dr Avisnata Das, Jincy Daniel, Anu Rose Andrews
29. Queen Elizabeth Hospital (Birmingham) - Prof Dhiraj Tripathi, Emma Burke, Emma Eaves, Helen Emms
30. Queen Elizabeth Hospital (Gateshead) - Dr Dina Mansour, Ann Wilson, Maureen Armstrong
31. Queen Elizabeth University Hospital (Glasgow) - Dr Rachael Swann, Faye McMeeken, Shona Perry
32. Queens Medical Centre (Nottingham) - Dr Naaventhan Palaniyappan, Elizabeth Davies, Kimberley Noon
33. Royal Berkshire Hospital - Dr Danielle Adebayo, Sarosh Khymani, Deepa Thapa
34. Royal Bolton Hospital - Dr Mahesh Bhalme, Emma McKenna, Julie Chadwick
35. Royal Bournemouth Hospital - Dr Jo Tod, Nina Barratt, Annamaria Wilce
36. Royal Derby Hospital - Dr Andrew Austin, Catherine Addleton
37. Royal Devon and Exeter Hospital - Dr Ben Hudson, Rob James, Lily Zitter, Jane Hall
38. Royal Free Hospital - Dr Jennifer Ryan, Christine Eastgate
39. Royal Liverpool University Hospital - Dr Edward Britton, Martina Lofthouse
40. Royal London Hospital (Barts) - Dr Vikram Sharma, James Hand, Louise Payaniandy, Paula Bravo
41. Royal Surrey County Hospital - Dr Marinos Pericleous, Sheila Mtuwa, Wisdom Mbama
42. Royal Sussex County Hospital - Dr Khaleel Jamil, Dr Sumita Verma, Dr Yaz Hassadin, Zhengmei He, Zdenka Cipinova
43. Royal Victoria Hospital - Dr Roger McCorry, Allison Lloyd, Heather Lawther
44. Southmead Hospital - Dr Zeino Zeino, Lana Ward, Trudie Burge
45. St George’s Hospital - Dr Sarah Hughes, Dr Joseph Delo, Criscel Jan Pelaez, David Whitley
46. St Mary’s Hospital - Dr Ameet Dhar, Dr Nowlan Selvapatt, Maria Lanoria
47. St Thomas’ Hospital - Dr Phil Berry, Dr Sreelakshmi Kotha, Jessica Cordle, Ankita Sunny
48. Sunderland Royal Hospital - Dr Rohit Sinha, Louise Fairlie, Jennifer Henderson
49. The James Cook University Hospital - Dr Darren Craig, Dr Eman Alabsawy, Julie Tregonning
50. Torbay Hospital - Dr Luke Summers, Sophy Booth
51. University Hospital Coventry - Dr Esther Unitt, Susan Dale
52. University Hospital of North Durham - Dr Francisco Porras Perez, Melanie Kent, Suzanne Naylor
53. University Hospital of Wales - Dr Tom Pembroke, Danielle Rice
54. Watford General Hospital - Dr Mohammed Shariff, Xiaobei Zhao
55. Wythenshawe Hospital - Dr Varinder Athwal, Alphonsa Biju, Sheetal Crasta

