## Supplementary material for "Beta-blockers or Placebo for Primary Prophylaxis (BOPPP) of oesophageal varices: Study protocol for a randomised controlled trial": SAP

**Beta-blockers Or Placebo for Primary Prophylaxis of oesophageal varices (BOPPP Trial). A blinded, UK multi-centre, clinical effectiveness and cost-effectiveness randomised controlled trial.**

**BOPPP Trial**

### Statistical Analysis Plan (SAP)

**Version 1.0 – 05/12/2023**

Version 0.1 started: 02/05/2019

Authors:

Professor Ben Carter, Dr Hassan Jafari

Based on BOPPP Protocol Version: 4.0, 31 MAY 2023

ISRCTN: ISRCTN10324656

This document presents statistical analysis plans up to date (05-12-2023), which are based on the most recent version of the protocol (BOPPP Protocol Version: 4.0, 31 MAY 2023). We hereby confirm that the entire process of revising the primary outcome in the protocol and updating the Statistical Analysis Plan was conducted while both statisticians were fully blinded to the trial arms. This rigorous adherence to blinding protocols ensures the objectivity and integrity of the statistical analysis and the impartiality of outcome assessment in this trial.

#### Purpose and Scope of Statistical Analysis Strategy

This document details the statistical analysis plan (SAP) for the primary paper reporting results from the BOPPP trial. The analysis will be presented in an analysis report, which will be used as the basis of the primary research publications according to the study publication plan. This SAP describes the statistical methods for the primary and secondary outcomes of the study as defined in the protocol. It is intended that the results reported in this paper will follow the strategy set out herein; subsequent papers of a more exploratory nature will not be bound by this strategy but will be expected to follow the broad principles laid down for the primary paper (or papers, should there be more than one). They are intended to establish the strategy that will be followed as closely as possible, when doing the main primary analysis and reporting of the trial. Reference was made to ICH guidelines on Statistical Principles [1] reference and CONSORT guidelines [2].

### Investigators:

##### Chief Investigator

Dr Vishal C. Patel, Chief Investigator
Consultant in Hepatology
Institute of Liver Studies, King's College Hospital NHS Foundation Trust
Denmark Hill, London, SE5 9RS


##### Chief Scientific Investigator

Dr Mark J. McPhail
Consultant in Hepatology & Intensive Care
Institute of Liver Studies, King's College Hospital NHS Foundation Trust
Denmark Hill, London, SE5 9RS


##### Senior Trial Statistician

Professor Ben Carter, Senior Statistician
Senior Lecturer in Biostatistics
Kings College London Clinical Trials Unit
PO64, Institute of Psychiatry, De Crespigny Park
Denmark Hill, London, SE5 9RF


##### Trial Statistician

Dr Hassan Jafari
Statistician, Department of Biostatistics and Health Informatics
Institute of Psychiatry, Psychology & Neuroscience at King's College London
De Crespigny Park, Denmark Hill, London, SE5 9RF


**SAP Sign off:**

DocuSigned by:

CI signature: 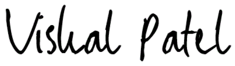

B4A414BC9C354AF...

Print Name: Dr. Vishal C. Patel

Date: 12/12/2023

Trial statistician signature: 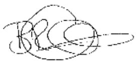

Print Name: Prof. Ben Carter

Date: **12/12/2023**

Independent DMEC/TSC statistician signature: 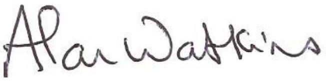

Print Name: Prof. Alan Watkins

Date: 8 December 2023

Chair of the TSC signature: 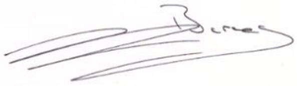

Print name: Prof. Ellie (Eleanor) Barnes

Date: 12th December 2023

#### List of Key Acronyms

| | |
|---|---|
| AE | Adverse Events |
| AUDIT-C | Alcohol Use Disorders Identification Test—Consumption |
| BOPPP | Beta-Blockers Or Placebo For Primary Prophylaxis Of Oesophageal Varices |
| CI | Confidence Interval |
| CLIF | Chronic Liver Failure Consortium Organ Failure Score |
| CONSORT | Consolidated Standards Of Reporting Trials |
| COPD | Chronic Obstructive Pulmonary Disease |
| CTIMP | Clinical Trial Of An Investigational Medicinal Product |
| CTO | Clinical Trial Organisation |
| DMC | Data Management Committee |
| DMEC | Data Management And Ethics Committee |
| DMP | Data Management Plan |
| MedDRA | Medical Dictionary For Regulatory Activities |
| EBL | Endoscopic Band Ligation |
| GCS | Glasgow Coma Scale |
| GI | Gastrointestinal |
| GOV2 | Gastroesophageal Varix - Type 2 |
| GP | General Practitioner |
| HCC | Hepatocellular Carcinoma |
| HE | Hepatic Encephalopathy |
| HES | Hospital Episode Statistics |
| HR | Hazard Ratio |
| ICH | International Council On Harmonisation Of Technical Requirements For Registration Of Pharmaceuticals For Human Use |
| IGV | Isolated Gastric Varices |
| IME | Important Medical Events |
| IMP | Investigational Medicinal Product |
| INR | International Normalised Ratio |
| ISRCTN | International Standard Randomised Controlled Trial Number |
| ITT | Intention To Treat |
| KCL | Kings College London |
| KCTU | Kings Clinical Trial Unit |
| KHP | Kings' Health Partners |
| LMM | Linear Mixed Model |
| MBOP | Mechanism Of Beta Blockade On Bacterial Translocation In Portal Hypertension |
| MDI | Multiple Deprivation Index |
| MELD | Model For End-Stage Liver Disease |
| MHRA | Medicines And Healthcare Products Regulatory Agency |
| NALFD | Non-Alcoholic Fatty Liver Disease |
| NHS | The National Health Service |
| NSBB | Non-Steroid Beta Blocker |
| OD | Omne In Die - Once Daily |
| OGD | Oesophagogastroduodenoscopy |
| OV | Oesophageal Varices |
| PD | Protocol Deviation |
| PIN | Participant Identification Number |
| PMN | Polymorphonuclear Neutrophils |
| PPI | Patients And Public Involvement |
| PPP | Per Protocol Population |
| PV | Protocol Violation |

| | |
|---|---|
| REC | Research Ethics Committee |
| SAE | Serious Adverse Event |
| SAP | Statistical Analysis Plan |
| SAR | Serious Adverse Reactions |
| SE | Standard Error |
| SOP | Standard Operating Procedures |
| SUSAR | Suspected Unexpected Serious Adverse Reaction |
| TIPS | Transhepatic Intrajugular Portosystemic Shunt |
| TMG | Trial Management Group |
| TSC | Trial Steering Committee |
| UKELD | United Kingdom Model For End-Stage Liver Disease |
| VH | Variceal Haemorrhage |

### Table of content


**1 Description of the trial ---- 12**

*1.1 Definition of all-cause decompensation ---- 13*

*1.2 Trial objectives ---- 13*

1.2.1 Primary objectives ---- 13

1.2.2 Secondary objectives ---- 13

*1.3 Trial endpoints ---- 14*

1.3.1 Primary endpoints ---- 14

1.3.2 Secondary & tertiary endpoints ---- 14

**2 Trial design ---- 15**

*2.1 Eligibility ---- 16*

*2.2 Method of allocation in groups ---- 17*

*2.3 Duration of the treatment and follow-up ---- 17*

*2.4 Frequency of follow-up ---- 17*

2.4.1 Visit windows ---- 17

*2.5 Trial Medication ---- 17*

2.5.1 Dosing Regimen ---- 18

2.5.2 Patient Permanent IMP discontinuation ---- 18

*2.6 Data collection ---- 20*

2.6.1 Measures: ---- 20

*2.7 Moderators and Mediators of treatment ---- 21*

*2.8 Sample size estimation ---- 21*

2.8.1 Estimation of all-cause decompensation rate ---- 21

2.8.2 Sample size calculation ---- 21

2.8.3 Interim analysis ---- 21

**3 Data analysis plan - Descriptive ---- 22**

*3.1 Recruitment and representativeness of recruited patients ---- 22*

*3.2 Population under investigation ---- 22*

*3.3 Baseline comparability of randomised groups ---- 22*

*3.4 Adherence to allocated treatment and treatment fidelity ---- 23*

3.4.1 Assessment of Prescription Adherence ---- 23

3.4.2 Reasons for Withdrawal ---------- 24
3.4.3 Treatment Compliance Analysis ---------- 24
3.4.4 Dose Modifications and Duration ---------- 24
3.4.5 Compliance Calculation ---------- 24
3.4.6 Adherence Formula ---------- 24
3.4.7 Adherent and Non-adherent Patients ---------- 24

*3.5 Loss to follow-up and missing data ---------- 25*
3.5.1 Missing baseline data ---------- 25
3.5.2 Missing items in scales and subscales (Questionnaires) ---------- 25
3.5.3 Missing time-to-event data ---------- 26
3.5.4 Missing IMP adherence data ---------- 26
3.5.5 Missing other outcome data ---------- 26
3.5.6 Missing Dates ---------- 27
3.5.7 General consideration on missingness ---------- 27

*3.6 Adverse event reporting ---------- 28*

*3.7 Concomitant Medications ---------- 29*

*3.8 Report of planned and unplanned unblinding ---------- 29*

*3.9 Descriptive statistics for outcome measures ---------- 30*
3.9.1 Laboratory Test Results ---------- 30
3.9.2 Vital Signs ---------- 30

**4 Data analysis plan – Inferential analysis ---------- 31**

*4.1 Main analysis of treatment differences ---------- 31*
4.1.1 Statistical considerations ---------- 31
4.1.2 Time of event (survival time) ---------- 31

*4.2 Analysis Datasets ---------- 31*
4.2.1 Intention-to-treat (ITT) population ---------- 31
4.2.2 Per protocol population (PPP) ---------- 31

*4.3 Data Screening, Cleaning and Acceptance ---------- 32*
4.3.1 General Principles ---------- 32
4.3.2 End of trial - database lock ---------- 32
4.3.3 Data Handling and Electronic Transfer of Data ---------- 33
4.3.4 Survival Data ---------- 33
4.3.5 Trial endpoints ---------- 33
4.3.6 Detection of Bias ---------- 34
4.3.7 Outliers ---------- 34
4.3.8 Statistical Assumptions and Distributional Characteristics ---------- 34
4.3.9 Statistical Analyses Quality control ---------- 34

*4.4 Planned Analyses* ---------- *35*

4.4.1 General Considerations ---------- 35

4.4.2 Interim Analysis and Early Stopping Guidelines ---------- 35

4.4.3 Analysis of primary outcome ---------- 36

4.4.4 Analysis of secondary outcomes ---------- 36

4.4.5 Statistical considerations ---------- 38

*4.5 Estimand Framework* ---------- *40*

4.5.1 Definition of Primary Estimand ---------- 40

4.5.2 Target Population: ---------- 40

4.5.3 Variable (or Endpoint): ---------- 40

4.5.4 Treatment or Intervention: ---------- 40

4.5.5 Population-Level Summary: ---------- 40

4.5.6 Post-Randomisation Events (Intercurrent Events): ---------- 40

4.5.7 Handling of Intercurrent Events in Analysis: ---------- 41

*4.6 Sensitivity analyses* ---------- *42*

4.6.1 Planned subgroup analyses ---------- 42

*4.7 Exploratory analyses* ---------- *43*

*4.8 Exploratory mediator and moderator analysis* ---------- *43*

**5 Software ---------- 43**

*5.1 Data management* ---------- *43*

*5.2 Statistical analysis:* ---------- *43*

**6 Reference list ---------- 43**

### 1 Description of the trial

Liver disease is the fifth commonest cause of death in the developed world and is rising in incidence, with liver failure a common mode of death in these patients.[1] The global disease burden of cirrhosis is rising owing to an increased prevalence of alcohol and non-alcoholic-related liver disease. In England and Wales, it is estimated that 60,000 people have cirrhosis with approximately 11,000 attributable deaths per annum [1][*]. Standardised mortality has risen by 400% from 1970, commonly in those of working age and in contrast to the other major causes of mortality.

Portal hypertension is a frequent clinical syndrome and complication of cirrhosis that is defined by an increase in porto-systemic pressure gradient in any portion of the portal venous system, and cirrhosis is the commonest cause of portal hypertension in 90% of cases. [2]

Of the complications that directly result from portal hypertension; the development of varices and variceal haemorrhage (VH) is one of the most significant. The management for varices and variceal haemorrhage has markedly advanced over the past decades. [3, 4]

Oesophageal varices (OV) are graded by size or features evident at oesophago-gastric duodenoscopy (OGD) consistent with increased risk of future haemorrhage (high risk stigmata). Grading of OV is by endoscopic appearance or estimation of diameter (≤5mm is deemed small) and directly affects the risk of VH or death.

Despite the advances of medical, endoscopic and radiological therapy the mortality rates of acute variceal haemorrhage is still 10%-20%.[5] Prevention of VH is therefore vital in those who have varices.

Prophylaxis against future haemorrhage can be by pharmacological methods or endoscopic therapies. The main pharmacological choice is non-selective beta-blockade (NSBB) and following several randomised controlled trials of different endoscopic methods band ligation is now the preferred endoscopic therapy of OV. The current evidence base, and international recommendations suggest that there is no benefit of NSBB in pre-primary prophylaxis for patients without varices. However, there is a clear benefit in the reduction of VH with NSBB in patients with moderate-large varices (>5mm in diameter), or those with advanced liver disease. [6] There is currently no clear evidence to guide the use of NSBB in patients with small varices.

Of the many NSBB used in clinical practice carvedilol has gained in favour over propranolol or other agents due to its dosing schedule, tolerability, and clinical effectiveness. While evidence is clarifying on the best method of primary prophylaxis in large varices [7] there is equipoise on whether those with small varices require primary prophylaxis at all. The implication for this project is that there is sufficient doubt to allow placebo as a control arm in comparisons of NSBB for primary prophylaxis in those with small varices and that carvedilol is an appropriate single agent to investigate.

[*] For references mentioned by number see trial protocol

The main benefits of NSBB include reduction in rate of progression to haemorrhage or progression to larger varices when initial variceal size is moderate [6] or more. This trial seeks to determine if there is a benefit for those with small varices. NSBBs are low cost, easy to administer and do not require specific expertise (they can be managed in primary care). As they act by decreasing portal pressure, NSBB may also reduce the development of ascites and decompensation. [8,9] Also, once a patient is on NSBB there may be no need for repeat Oesophago-gastric duodenoscopy (OGD).

Therefore, variceal haemorrhage is the primary reason for the potential use of NSBB in this cohort of patients. NSBB are hoped to prevent progression of portal hypertension to the point where VH occurs and thus prevent hospital admission and the risk of death associated with VH. The PPI group confirmed this as a patient supported preferred outcome.

#### 1.1 Definition of all-cause decompensation

All-cause decompensation is defined within BOPPP as the occurrence of any of the below:

- Variceal haemorrhage
- New or worsening ascites
  - Defined by clinical examination or radiological findings.
- New or worsening hepatic encephalopathy.
  - Defined by West-Haven Grade >1 (overt HE)
- Spontaneous bacterial peritonitis
  - Ascitic fluid cell PMN cell count >250/mm3
- Hepatorenal syndrome
- Increase in Child Pugh Grade by one grade or MELD by five points (Appendix 5)
- Liver-related mortality

#### 1.2 Trial objectives

The aim of this study is to evaluate the clinical efficacy and cost-effectiveness of NSBB in reducing disease progression, defined by all-cause decompensation, in cirrhotic patients with small varices.

##### 1.2.1 Primary objectives

• To determine the clinical effectiveness in the reduction of all-cause decompensation in patients treated with carvedilol versus placebo up to 3 years.

• To determine the cost-effectiveness of Carvedilol in patients with small oesophageal varices. (The cost-effectiveness will be analysed and reported by Health Economist team, and the methods and plan are out of the scope of this SAP).

##### 1.2.2 Secondary objectives

- At 1-year after participant recruitment opens, to assess feasibility of: recruitment, retention acceptability, with progression criteria outlined in internal pilot I.
- To determine additional clinical benefits of Carvedilol versus placebo for: reduction of variceal size progression, need to initiate endoscopic management of varices (endoscopic band ligation (EBL)), deterioration in liver function (assessed by MELD score and Child-Pugh grade) and all-cause mortality.
- To investigate how this is best delivered in primary care, by general practice, using qualitative approaches and GP interviews to examine barriers and enablers to implementation.

#### 1.3 Trial endpoints

##### 1.3.1 Primary endpoints

1. Time to first decompensating event.
2. Cost-utility of NSBB over trial follow-up to 3 years.

##### 1.3.2 Secondary & tertiary endpoints

1. Estimation of the one, and 3-year variceal haemorrhage rate by allocation, and associated number needed to treat.
2. Progression to medium/large oesophageal varices at gastroscopy requiring clinical intervention.
3. Development of gastric, duodenal, or ectopic or rectal varices in the GI tract at gastroscopy
4. Composite of progression in variceal size and haemorrhage as per 1 and 2 by 3 years.
5. Survival (Overall, liver-related, cardiovascular-related).
6. Quality of life, EQ-5D-5L.

All the above will be analysed until 3 years from randomisation. We will seek permission for collection of primary outcome and Health Episode Statistic (HES) data post cessation of IMP until trial completion.

### 2 Trial design

A UK wide, multicentre, Phase IV, blinded (participant, outcome assessor, investigator, chief investigators, and senior statistician), randomised controlled trial of beta blockade with Carvedilol versus Placebo in patients with cirrhosis and small varices without evidence of previous haemorrhage in England, Wales, Scotland and Northern Ireland.

This trial will incorporate both a clinical primary outcome and cost effectiveness outcome to evaluate the benefit to patients and society for earlier intervention with beta-blockers. The clinical primary endpoint is time to all-cause decompensation from baseline until 3 years (with further data collection until last patient last visit).Figure 1 represents the flow diagram for the trial BOPPP.

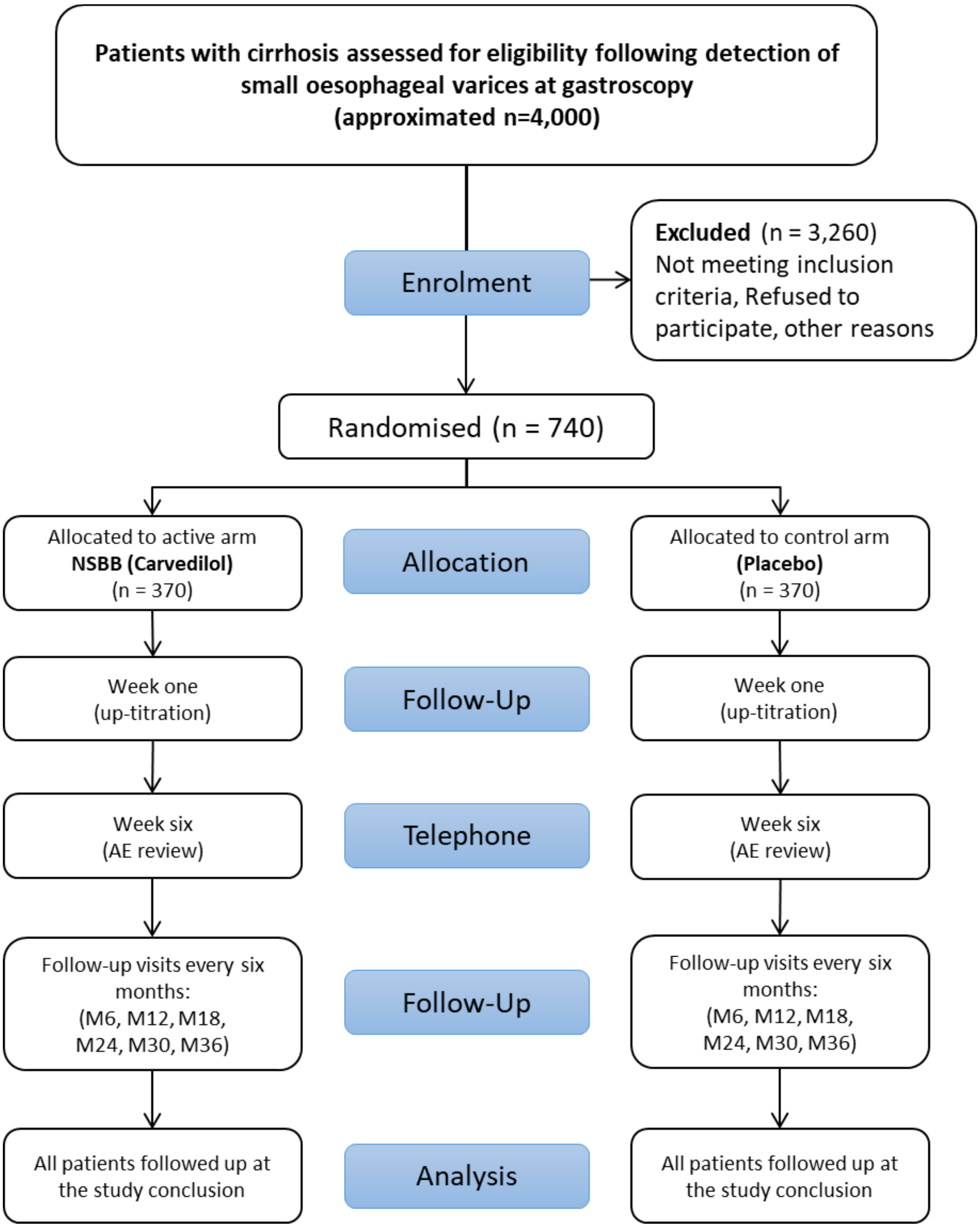


**Figure 1. Trial design flow diagram**

#### 2.1 Eligibility

The eligibility criteria are listed in the **Table 1**.

**Table 1. list of eligibility criteria**

| Inclusion: |
|---|
| 1. Age 18 years and over |
| 2. Cirrhosis and portal hypertension, defined by any two of the following: |
| • Characteristic clinical examination findings; one or more of |
| • Characteristic liver function tests |
| • Haematological panel |
| • Coagulation profile abnormalities |
| • Characteristic radiological findings; one or more of |
| • Heterogeneous liver with irregular contour |
| • Splenomegaly |
| • Ascites |
| • Varices |
| • Recanalized umbilical vein |
| • FibroScan liver stiffness measurement >15 kPa without other explanation |
| • Fibrosis score > ISHAK stage 4 on liver biopsy (presence of a relevant fibrosis score by biopsy does not require additional clinical examination / radiological / FibroScan supporting evidence) |
| 3. Small oesophageal varices diagnosed within the last 6 months |
| 4. Not received a beta-blocker in the last week |
| 5. Capacity to provide informed consent |
| **Exclusion:** |
| 1. Non-cirrhotic portal hypertension |
| 2. Current medium/large oesophageal varices (defined as >5 mm in diameter) |
| 3. Previous medium/large oesophageal varices (defined as >5 mm in diameter), which decreased in size with curative therapy |
| Gastric (IGV and GOV2), duodenal, rectal or other ectopic varices with or without evidence of recent haemorrhage. For gastric varices, this includes: |
| • IGV-1 and IGV-2 (isolated gastric varices) |
| • GOV2 (gastric varices continuing into the cardia) |
| • Note GOV1 (gastric varices continuing into the lesser curve) are not an exclusion if present with small oesophageal varices |
| 4. Previous variceal haemorrhage |
| 5. Previous band ligation or glue injection of oesophageal and/or gastric varices |
| 6. Red signs accompanying varices at endoscopy |
| 7. Known intolerance to beta blockers |
| 8. Contraindications to beta blocker use: |
| • Heart rate <50 bpm |
| • Known second degree or higher heart block |
| • Sick sinus syndrome |
| • Systolic blood pressure <85 mmHg |
| • Chronic airways obstruction (asthma/COPD) |
| • Floppy Iris Syndrome |
| • CYP2D6 Poor Metaboliser |
| • History of cardiogenic shock |
| • History of severe hypersensitivity reaction to beta-blockers |
| • Untreated phaeochromocytoma |
| • Severe peripheral vascular disease |
| • Prinzmetal angina |
| • NYHA IV heart failure |
| 9. Unable to provide informed consent |
| 10. Child Pugh C cirrhosis |
| 11. Already receiving a beta-blocker for another reason that cannot be discontinued |
| 12. Graft cirrhosis post liver transplantation |
| 13. Evidence of active malignancy without curative therapy planned |
| 14. Pregnant or lactating women |
| 15. Women of childbearing potential not willing to use adequate contraception during the period of IMP dosing* |
| 16. Patients who have been on a CTIMP within the previous 3 months |
| 17. Clinical symptoms consistent with COVID-19 (a high temperature, a new continuous cough or a loss or change to sense of smell or taste) at the time of randomisation |

#### 2.2 Method of allocation in groups

Once baseline assessments are complete and following consent has been obtained, the individuals will be randomised to one of the treatment arms (Carvedilol versus Placebo). Randomisation will be done in a 1:1 ratio. Randomisation is at the patient level in each site and is performed using an online randomisation system set up by the King's Clinical Trials Unit (KCTU) at the Institute of Psychiatry, Psychology & Neuroscience in London. Randomisation is stratified by site to ensure that equal numbers of patients are allocated to the treatment arms within each site, with variable block sizes. The procedure is as follows: On receipt of the baseline questionnaire, the Research Nurse electronically submits details of each participant to the KCTU Randomisation system. This includes participant ID number (PIN), site name, initials, and date of birth. The system immediately notifies the relevant trial team members and records the randomisation outcome.

#### 2.3 Duration of the treatment and follow-up

The duration of the treatment will be 3 years (36 months) after post randomisation for each participant. To answer the trial question in an ethical, pragmatic, and cost-effective manner we have set a combined 3-year follow up or a minimum number of decompensation events (n=185) follow up. Thereafter we would seek permission to obtain further information on certain outcomes after the patients' last visit (e.g., at 36 months); i.e., haemorrhage; progression; mortality; health care utilisation as part of a post IMP follow-up period. This will increase the power of the trial and add important additional weight to the findings without unduly impacting on the trial budget or burden to the patient. Future access to patient records will be explicitly requested from patients at the time of consent.

#### 2.4 Frequency of follow-up

When the participant enrols to the trial there will be a screening and baseline visit which will happen in a single session typically. Participants will complete follow up measures at every 6 months for up to 3 years (or after minimum of 185 events). Follow-Up visits will be at 1 week and at 6, 12, 18, 24, 30, and 36 months after randomisation. The 1-week visit will be for up-titration of the drug. In this visit only the vital signs and IMP dose log data will be collected.

##### 2.4.1 Visit windows

The visit windows will be +/- 3 days for the week 1 visit and +/- 6 weeks for month 6, 12, 18, 24, and 36 visits.

#### 2.5 Trial Medication

The trial involves two arms: Carvedilol tablets (active IMP) and matching placebo tablets. Carvedilol, a vasodilatory non-selective beta-blocker, is presented as oval, white tablets and used for reducing heart rate and peripheral vascular resistance. The starting dose is 6.25 mg once daily (OD), with a possible increase to 12.5 mg OD based on tolerability, aiming for a target heart rate of 50-55 bpm. For cirrhosis patients, 12.5 mg OD is the expected maximum tolerable dose. Placebo tablets, identical in appearance to Carvedilol, are also used for blinding.

##### 2.5.1 Dosing Regimen

Participants will start with 6.25 mg OD of Carvedilol or placebo, potentially increasing to 12.5 mg OD after one week. Dose adjustments may occur based on clinician discretion and patient side effects. The trial aims for a 25% reduction in baseline heart rate, targeting 50-55 bpm. Carvedilol is expected to be well-tolerated with a low rate of compliance issues or discontinuation due to adverse events (AEs). The trial medication will be dispensed every six months for three years, with dose management and follow-up incorporated at various stages, including initial escalation, a week six telephone call, and subsequent 6-month intervals.

##### 2.5.2 Patient Permanent IMP discontinuation

Participants can withdraw from the trial medication at any time. Investigators also have the discretion to withdraw participants due to various reasons including inter-current illnesses, adverse events (AEs), serious adverse events (SAEs), suspected unexpected serious adverse reactions (SUSARs), repeated protocol deviations/violations, cure, or other specific criteria. Notable withdrawal criteria include the need for beta-blockade following a cardiac or cerebrovascular event, and withdrawal of patient consent.

If withdrawal from the trial medication is necessary, detailed reasons will be documented. Unless consent for follow-up is withdrawn, these participants will continue in the trial, and efforts will be made to collect safety and endpoint data. This includes follow-up until the last patient visit, even if the IMP is discontinued before the end of the 3-year trial period. Data collection will encompass laboratory parameters, quality of life, healthcare utilisation, and alcohol consumption data.

Participants should be permanently withdrawn from the trial medication if they experience events indicative of disease progression, such as an increase in oesophageal varices to grade II/III, development of varices in other GI tract locations, or undergo interventions like a Transjugular Intrahepatic Porto-Systemic Shunt (TIPS) or liver transplantation. Standard of Care (SoC) will be provided, which may include commencement of non-selective beta-blockers (NSBB) and/or endoscopic band ligation. For such participants, IMP will be discontinued, and they will be withdrawn from further 6-monthly follow-up, although mortality and endpoint data will still be collected at the trial's end. See Figure below for an illustrative diagram of the process for temporary and permanent discontinuation of the IMP.

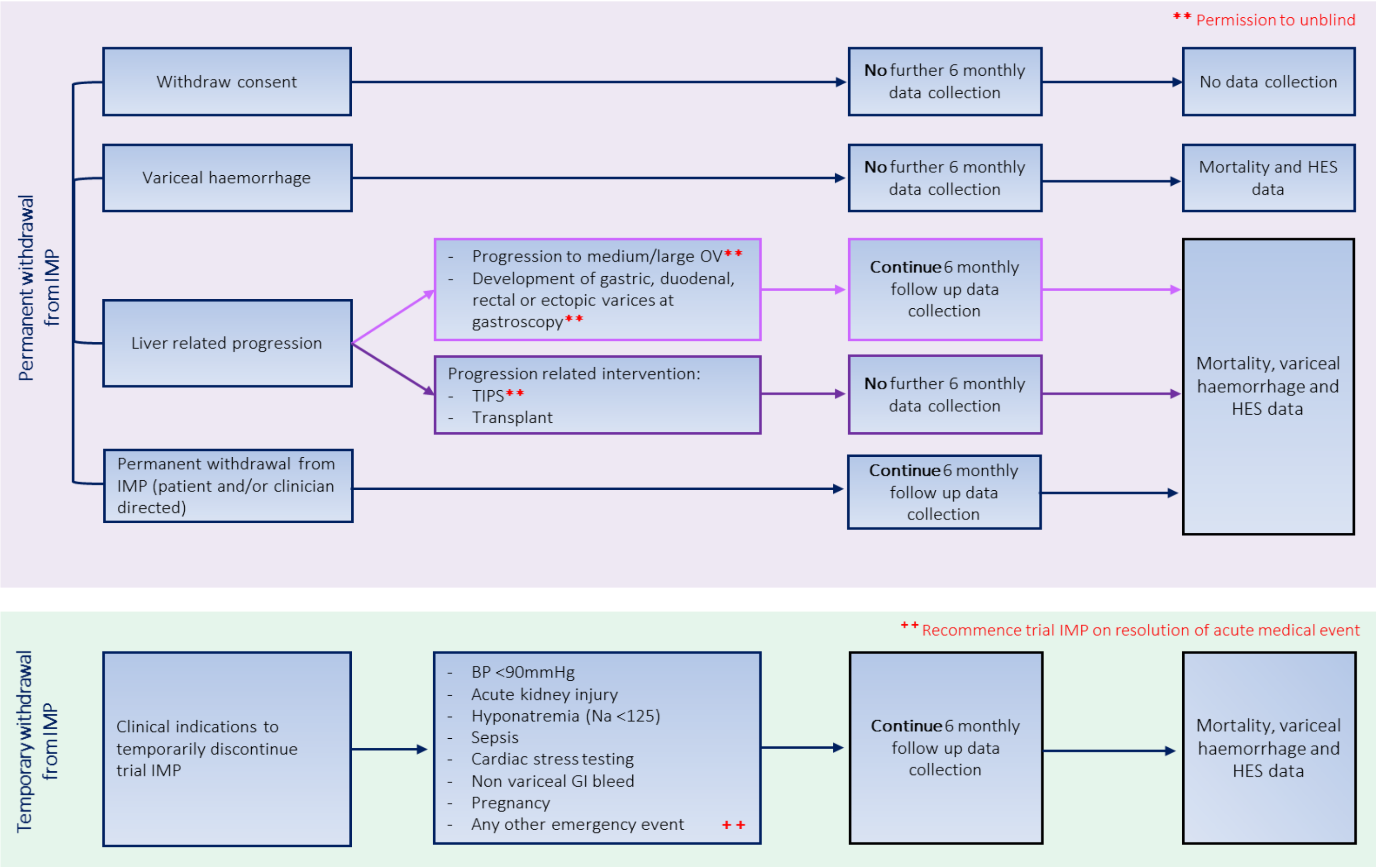


**Figure 2. Follow-up schedule pertaining to events leading onto permanent or temporary withdrawal from trial IMP.**

#### 2.6 Data collection

##### 2.6.1 Measures:

**Table 2. list of Baseline, Week 1, Week 6, 6 monthly Follow-Ups, and Continuous Data Collection variables.**

| Variables | B | W 1 | W 6 | M 6 | M 12 | M 18 | M 24 | M 30 | M 36 | At VH* |
|---|---|---|---|---|---|---|---|---|---|---|
| **Demographics** | | | | | | | | | | |
| Age | • | | | | | | | | | |
| sex | • | | | | | | | | | |
| Ethnicity | • | | | | | | | | | |
| Smoking/Vape status | • | | | | | | | | | |
| Recreational drug use | • | | | | | | | | | |
| Highest level of education | • | | | | | | | | | |
| Employment status | • | | | | | | | | | |
| Multiple Deprivation Score Index (MDI) | • | | | | | | | | | |
| Height | • | | | | | | | | | |
| **Medical History** | • | | | | | | | | | |
| **Cirrhosis History (Primary and secondary cause)** | • | | | | | | | | | |
| **Alcohol consumption /AUDIT-C** | • | | | • | • | • | • | • | • | |
| **Smoking status** | • | | | • | • | • | • | • | • | |
| **Targeted physical exam** | | | | | | | | | | |
| Grade of hepatic encephalopathy* | • | | | • | • | • | • | • | • | • |
| Glasgow Coma Scale Score (GCS) | • | | | • | • | • | • | • | • | • |
| Ascites* | • | | | • | • | • | • | • | • | • |
| Weight (kg) | • | | | • | • | • | • | • | • | • |
| Dialysis History | • | | | • | • | • | • | • | • | • |
| Hepatocellular carcinoma (HCC) – in 6 months ago | • | | | • | • | • | • | • | • | • |
| **Gastroscopy (collection at every one-year visit)** | | | | | | | | | | |
| Number of columns of oesophageal varices (grade I, II, III) | • | | | | • | | • | | • | • |
| Oesophageal varices: presence of red signs | • | | | | • | | • | | • | • |
| Gastric varices presence: (GOV1, GOV2, IGV1, IGV2) | • | | | | • | | • | | • | • |
| Duodenal varices presence | • | | | | • | | • | | • | • |
| Endoscopic Band Ligation (EBL) | • | | | | • | | • | | • | • |
| **HCC surveillance ultrasound (US)** | | | | | | | | | | |
| Hepatomegaly | • | | | • | • | • | • | • | • | • |
| Splenomegaly | • | | | • | • | • | • | • | • | • |
| Spleen size | • | | | • | • | • | • | • | • | • |
| Hepatic vein patency | • | | | • | • | • | • | • | • | • |
| Portal vein patency | • | | | • | • | • | • | • | • | • |
| Radiological ascites | • | | | • | • | • | • | • | • | • |
| Focal liver lesions | • | | | • | • | • | • | • | • | • |
| **Vital signs** | | | | | | | | | | |
| Systolic blood pressure | • | • | | • | • | • | • | • | • | • |
| Diastolic blood pressure | • | • | | • | • | • | • | • | • | • |
| Heart Rate | • | • | | • | • | • | • | • | • | • |
| **Child Pugh score*** | • | | | • | • | • | • | • | • | • |
| **MELD (Model End Stage Liver Disease) score*** | • | | | • | • | • | • | • | • | • |
| **UKELD (United Kingdom Model for End-Stage Liver Disease) score** | • | | | • | • | • | • | • | • | • |
| **CLIF-C-AD score** | • | | | • | • | • | • | • | • | • |
| **Blood test†** | • | | | • | • | • | • | • | • | • |
| **Quality of Life - EQ-5D-5L** | • | | | • | • | • | • | • | • | |
| **Resource use** | • | | | • | • | • | • | • | • | |
| **Telephone call only** | | | • | | | | | | | |
| **Concomitant Medications** | | • | • | • | • | • | • | • | • | |
| **IMP Adherence log** | | • | • | • | • | • | • | • | • | |
| **IMP dose Log** | | • | • | • | • | • | • | • | • | |
| **Adverse Event Log*** | | • | • | • | • | • | • | • | • | |
| **VH event occurrence*** | | • | • | • | • | • | • | • | • | |
| **Withdrawals and Mortality*** | | • | • | • | • | • | • | • | • | |

† Blood test encompasses Full Blood Count (including White blood cell count, Neutrophils, Lymphocytes, Monocytes, Eosinophils, Basophils, Haemoglobin, Platelet count, Mean cell volume), International Normalised Ratio (INR), Liver Profile (Alanine transferase, Aspartate transaminase, Total bilirubin, Alkaline phosphatase, Albumin, Gamma glutamyl transferase, Total protein), Renal Profile (Sodium, Potassium, Creatinine, Urea), Bone profile (corrected calcium), and Glucose. * Items marked with an asterisk (*) represent the elements constituting the primary outcome of the study. B: Baseline, W: Week, M: Month

#### 2.7 Moderators and Mediators of treatment

This SAP outlines that no formal mediation or moderation analyses are explicitly planned as part of the primary objectives of the study. In the context of ancillary analyses, there may be an exploratory investigation into the moderation of the treatment effect, contingent upon the identification of significant covariates. Furthermore, adjustments or controls for the impact of potential moderating variables may be considered in the analysis as deemed appropriate. The mediation Mechanistic analysis will be formally explored within the scope of the **MBOP** sub-study. This analysis will be conducted to investigate the underlying mechanisms and pathways through which the treatment effects are exerted, providing deeper insights into the causal relationships within the study.

#### 2.8 Sample size estimation

##### 2.8.1 Estimation of all-cause decompensation rate

The PREDESCI study enrolled 201 patients (1:1) and reported 27%, versus 16% 3-year decompensation rate in the placebo and β-blockers group (HR 0·51, 95% CI 0·26–0·97, p=0.04). A BOPPP blinded data extract in March 2022 showed that our all-cause decompensation rate was 20% at 1-year (across both arms combined).

##### 2.8.2 Sample size calculation

Based on the data collected from 187 patients randomised in the BOPPP trial as of December 2021, the event rate of all-cause decompensation in both study arms was approximately 0.16 at one year. To ensure a conservative estimate and account for possible censoring due to intercurrent events, such as adverse events that may cause treatment discontinuation in these patients, we have projected that the 3-year decompensation rate in the BOPPP trial will be at least 25.5%, or 31% in the placebo arm and 20% in the β-blocker arm. To detect this difference with a hazard ratio of 0.60, we require a total of 666 patients (or 170 events) to achieve 90% power and a type-1 error rate of 0.05. To account for a potential 10% loss to follow-up, we plan to enrol 740 patients. Should the true hazard ratio be 0.70, and the event rate in the control arm remains the same, the trial will retain 71% power.

##### 2.8.3 Interim analysis

No interim analysis is planned for efficacy or event rate, beyond the feasibility pilot phase assessments.

###### 2.8.3.1 Internal pilot (Recruitment and retention)

An internal pilot is planned to determine the feasibility of the trial and will be run at 12 months after recruitment opens, with the following Go/No Go progression to demonstrate the ability of recruit sites and patients within the sites:

1. To have opened at least eight sites, with at least one patient randomised at each.
2. To have randomised at least 80 patients.
3. To have a retention rate of at least 70%.

If the above criteria are met, we will progress the trial.

### 3 Data analysis plan - Descriptive

#### 3.1 Recruitment and representativeness of recruited patients

CONSORT flow chart will be constructed – see **Figure 3. CONSORT Flow Diagram** for template. This will include the number of screened patients, eligible patients, number of patients agreeing to enter the trial, number of exclusions, number of patients refusing, or missed to follow-up for any other reason, then by treatment arm (the number continuing through the trial, the number withdrawing, the number lost to follow-up and the numbers analysed). The number and percentages will be presented for each treatment arm and for each follow-up visits.

If the patient has a variceal haemorrhage, they should discontinue IMP, and should be started on NSBB as per clinical standard of care. Similarly, if during surveillance (planned or unplanned), the patient's varices have progressed to medium/large then they also require band ligation and/or NSBB and they will discontinue the IMP and the planned visits and be started on NSBB. The patients who experience the variceal haemorrhage will be traced for mortality, and the patients who progress will be traced for variceal haemorrhage and mortality through medical and hospital reports at the end of the trial.

#### 3.2 Population under investigation

An intention to treat (ITT) population will encompass all patients involved in the study's outcome assessment. Patients who encounter any of the all-cause decompensation events listed in section 1.1, whichever occurs first, will be classified as having treatment failure and will be recorded as an event. Patients who do not undergo any form of decompensation during the follow-up period will be considered non-events and censored at the last time of follow-up time.

The per-protocol population (PPP) excludes participants who violates the study protocol. In the analysis of the primary outcome, participants who progress (e.g., due to an increase in varices size), but have not experienced any type of decompensation will be censored.

#### 3.3 Baseline comparability of randomised groups

The assessment of comparability includes pertinent demographics and socioeconomic characteristics, risk of prognostic factors, and medical history. The investigator and statisticians can only look at the relevant factors at baseline about which they were aware and were measured. Obviously, those which are unknown cannot be compared. Randomisation on average produces balance between comparison groups, clearly imbalances are common in smaller trials and would be unlikely in BOPPP trial. The evaluation of treatment comparability between groups, (carvedilol) and control, will include demographics, baseline disease characteristics, and medical history. Descriptive statistics (mean, standard deviation, median, range) will be presented by treatment group for continuous variables. Categorical variables will be summarised using frequencies and percentages by each arm. The baseline differences between the two randomised groups will not be tested for significance, however presented descriptively. According to the CONSORT statement, significance testing of baseline differences in randomised controlled trials should not be performed. (Moher et al.,

2010), unless otherwise specified. The baseline value listed in Table 2 will be defined as the closest measurement prior to the first dose of study IMP, in case of multiple record.

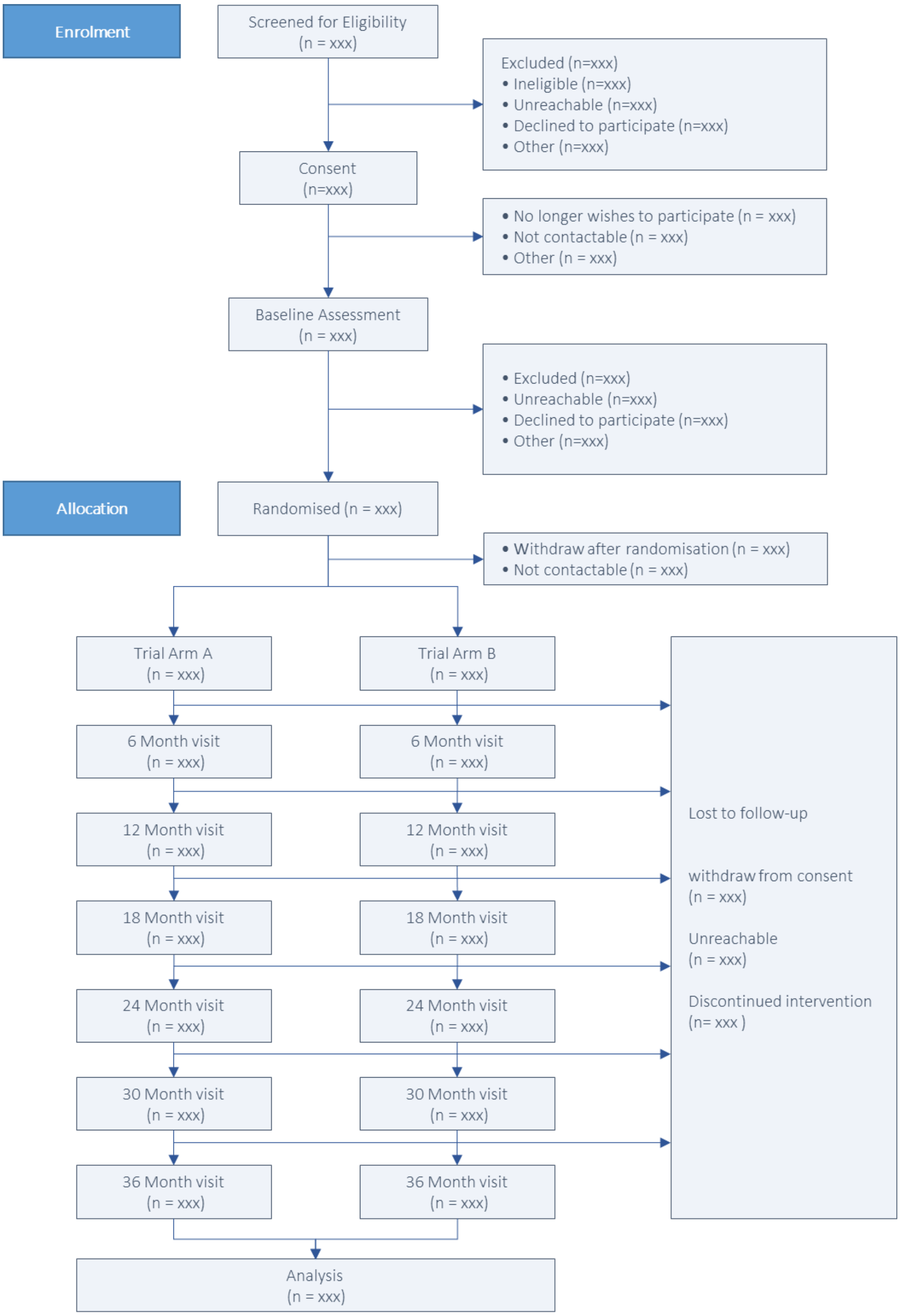


**Figure 3. CONSORT Flow Diagram**

#### 3.4 Adherence to allocated treatment and treatment fidelity

##### 3.4.1 Assessment of Prescription Adherence

The frequencies and proportions of participants who received a prescription for their allocated treatment at randomisation will be detailed both graphically and in tabular format, for each randomised group and overall.

Additionally, the proportion of participants who discontinue their IMP (withdraw from IMP) will be summarised by randomised group.

##### 3.4.2 Reasons for Withdrawal

The reasons for withdrawal from the IMP will be collated and summarised.

##### 3.4.3 Treatment Compliance Analysis

The primary population for assessing treatment compliance includes all patients who receive at least one prescription of the trial IMP. Compliance and non-compliance will be evaluated in relation to baseline variables, and allocated treatment arm.

##### 3.4.4 Dose Modifications and Duration

Modifications in dosage will be described, and treatment duration will be defined as the period from the first prescription of the trial drug to the last dose taken by the patient. The total dose taken will be calculated as 6.25 mg IMP multiplied by the number of tablets consumed, determined by subtracting the number of returned tablets from those dispensed at each visit. The expected total dose for each patient will be the assigned daily dose multiplied by the total number of tablets dispensed during the treatment duration.

##### 3.4.5 Compliance Calculation

Overall IMP compliance will be calculated as the total dose taken during the trial divided by the expected total dose, expressed as a percentage. Generally, each patient is expected to take a maximum of two 6.25 mg tablets per day. In cases of dose reductions, the expected number of tablets will reflect the new, lower dosage. Dose reductions or interruptions will be summarized by treatment group.

##### 3.4.6 Adherence Formula

Adherence will be quantified using the formula:

$$Adherence = \frac{Number\ of\ dosage\ units\ dispensed - \ number\ of\ dosage\ units\ remained}{prescribed\ number\ of\ dosage\ unit\ per\ day\ \times\ number\ of\ days\ between\ 2\ visits} \times 100$$

**Example Calculation:** For a patient prescribed 12.5 mg of IMP per day who receives two bottles (each containing 210 tablets) and returns 80 tablets after 180 days, the adherence would be calculated as:

$$Adherence = \frac{420 - \ 80}{2\ \times\ 180} \times 100\ =\ 94\%$$

##### 3.4.7 Adherent and Non-adherent Patients

**Defining Adherence Levels:** The adherence value, as measured by the IMP adherence log, will be used to categorise patients into groups of good and poor compliance. A threshold of 50% adherence has been established as a reasonable cut-off point to differentiate between adherent and non-adherent patients, in line with the trial protocol.

**Subgroup Analysis:** Subgroup analyses will be conducted to compare the outcomes of patients who are fully adherent to the treatment regimen ('adherent') with those who are partially adherent ('non-adherent'). This analysis will be performed if deemed appropriate based on the trial data.

For missingness in IMP adherence and management log please see section 3.5.4.

#### 3.5 Loss to follow-up and missing data

**Patient Flow Reporting:** The flow of patients through the trial will be comprehensively documented. This includes the number of patients screened, meeting the inclusion criteria, randomised, eligible but not randomised, withdrawing post-randomisation, and lost to follow-up. These details will be illustrated in CONSORT flow diagrams, as depicted in **Figure 3.**

**Discontinuation and Follow-Up Data:** The number and percentage of participants discontinuing the trial will be tabulated by specific reasons for discontinuation. Additionally, patient attendance at scheduled follow-up visits (conducted every 6 months up to 3 years) will be reported, both numerically and as a percentage.

**Missing Data on Outcomes:** The proportions of participants with missing data for primary and main secondary outcome measures will be summarised for each treatment arm and at each relevant time point. The reasons for withdrawal from the trial, along with corresponding data, will be summarised and presented in Table 3.

**Table 3. Reasons for Withdrawal from Trial by Treatment Arm**

| Reason for withdrawal | Treatment | Placebo |
|---|---|---|
| Participant no longer wishes to have further data collected | xxx (%) | xxx (%) |
| Participant unable to be located/contacted | xxx (%) | xxx (%) |
| Participant unable to travel to centre | xxx (%) | xxx (%) |
| Death of participant | xxx (%) | xxx (%) |
| Adverse event | xxx (%) | xxx (%) |
| Other | xxx (%) | xxx (%) |

##### 3.5.1 Missing baseline data

We anticipate minimal missing baseline data in this trial. Where missing baseline data exists, and modelling requires the use of a baseline measure we will impute using a method suitable to the variable as per the recommendations of White and Thompson. (White & Thompson, 2005)

##### 3.5.2 Missing items in scales and subscales (Questionnaires)

**Reporting Complete Data:** The proportion of participants with complete data for each scale and subscale in questionnaires will be reported as a percentage. **Handling Missing Data in Questionnaires:** In cases where specific guidelines for handling missing data in a questionnaire are available, these guidelines will be followed. If the primary source of a questionnaire does not provide instructions for managing missing data, a pro-rata scoring approach will be employed. This method involves replacing missing items with the mean of the non-missing items, on the condition that the level of missingness is less than 20%. Therefore, a total score will be calculated as long as at least 80% of the questionnaire items are completed.

##### 3.5.3 Missing time-to-event data

**Determining Event-Free Status:** For the primary outcome and other time-to-event data, the patient's status will be determined using various forms and medical records, including the Status Form, Withdrawal Form, Gastroscopy Form, Trial Completion Review Form, Targeted Physical Exam Form, MELD Score and Child Pugh Forms, Adverse Events Logs, Variceal Haemorrhage Form, and the patient's Medical Notes. If these sources do not report any relevant events, the patient will be classified as event-free at the study's end.

**Handling Missed Visits and Loss to Follow-Up:** In cases where a patient misses a visit or is lost to follow-up, their status regarding primary and secondary endpoints will be determined based on subsequent visit data or medical notes. If there is no report of an event at the trial's conclusion (as per patient notes), it will be assumed that the event did not occur.

**Censoring Procedure:** Patients with no reported events will be censored at the last known date they were tracked, either through any of the mentioned forms or medical notes. This ensures that their data is included up to the point of last known status.

##### 3.5.4 Missing IMP adherence data

**Tracking Incomplete Treatment:** The number and percentage of patients experiencing interruptions in IMP (Investigational Medicinal Product) treatment will be summarized by treatment arm. This includes cases where patients discontinue the intervention but continue with data collection.

**Subgroup Analysis:** If appropriate, subgroup analyses will be conducted comparing patients with complete (adherent) versus partial (non-adherent) treatment courses.

###### 3.5.4.1 Handling Missing Data on IMP Administration:

**1.** If a Patient Forgets to Bring the Medication Bottle: In cases where patients fail to bring their medication bottle to a visit, the number of tablets taken will be estimated based on their response in the IMP Adherence Log Form (Question ADH_02: "How often did the participant take the tablets on average since the last visit?"). The responses 'Always', 'Very Often', 'About Half the Time', 'Rarely', and 'Never' will correspond to adherence rates of 100%, 85%, 50%, 25%, and 0%, respectively.

**2.** If a Patient is Lost to Follow-Up: For patients who become lost to follow-up, the IMP adherence for their last 6-month interval with the received drug will be estimated using the mean adherence values from their previous intervals.

##### 3.5.5 Missing other outcome data

###### 3.5.5.1 Handling Continuous Repeated Measures:

<u>Approach:</u> For continuous outcomes measured repeatedly, missing post-randomisation assessments will be addressed by using mixed models fitted to all available data, employing maximum likelihood methods. <u>Assumptions:</u> This approach assumes an ignorable (or Missing at Random - MAR) missing data mechanism, which is a standard assumption in such

analyses (referenced: Little & Rubin, 2002; Molenberghs & Kenward, 2007).

Ensuring MAR Validity: To support the MAR assumption, we will investigate if baseline variables are predictive of missing outcome data at different time points. Variables found to be predictive will be included in the models to adjust for potential biases.

###### 3.5.5.2 Handling Binary Secondary Outcome (Variceal Size Progression):

Likelihood of Missingness: Missing data for the binary secondary outcome (progression of variceal size) is expected to be minimal, as this information is typically obtained from annual gastroscopy reports.

Resolution of Missing Data: In cases of missing data, queries will be sent to the respective sites for resolution. Missing data at one time point will be imputed using data from the subsequent time point. For instance, if variceal progression data is missing for the 12-month visit, it will be imputed with data from the 24-month visit, or, if not available, from the 36-month visit.

##### 3.5.6 Missing Dates

Initial Inquiry and Data Cleaning:

Process for Partially or Completely Missing Dates: In cases where dates for any primary or secondary events are partially or completely missing, these gaps will be addressed during the data cleaning phase. An initial inquiry will be made to correct or complete the missing date information. Estimation of Dates (post-inquiry procedure): If the precise dates remain unknown even after inquiry, the Trial Management Group (TMG) will estimate the most probable date of the event. This estimation will be based on a comprehensive review of the available medical notes and other relevant documentation. Rationale for Estimation: This approach is taken to ensure that the most accurate timeline of events is established, which is crucial for the integrity of the trial's analysis.

Handling Complete Absence of Date Information:

Censoring Procedure: In situations where there is no indication of any events in the forms or medical notes, and the date cannot be estimated, the patient will be censored at the last known date they were tracked in the medical notes. Importance of Censoring: Censoring at the last known date maintain the integrity of the analysis, ensuring that each participant's data contributes up to the point of their last confirmed status.

##### 3.5.7 General consideration on missingness

All of the above methods assume that missing values are missing at random, and unbiased results can be obtained using survival analysis. This assumption, however, may not be appropriate in some instances. (Little & Rubin, 2002; Molenberghs & Kenward, 2007). The baseline characteristics of those missing follow up will be compared to those with complete follow up.

#### 3.6 Adverse event reporting

We are recording Adverse Events (AEs) and recording and reporting serious adverse events (SAEs) in this trial. Participants are asked about any AEs at every consultation and AEs can be recorded ongoing through the trial. All serious adverse events and important medical events (IME) will be reported to the sponsor King's Health Partners Clinical Trial Office (KHP-CTO) who will report to the MHRA and REC. The following information mentioned in Table 4 and Table 5 will be recorded and reported.

The number of serious adverse events (SAE), serious adverse reactions (SAR) and serious unexpected adverse reactions (SUSAR) will be summarised overall and by treatment group as number of events and number of people who experienced events.

**Table 4. Adverse events frequency by body system code**

| Adverse Event by | Treatment | | Placebo | |
|---|---|---|---|---|
| Body system code | Participant | Incident | Participant | Incident |
| Infections and infestations | n (%) | n (%) | n (%) | n (%) |
| Blood and lymphatic system disorders | n (%) | n (%) | n (%) | n (%) |
| Vascular disorders | n (%) | n (%) | n (%) | n (%) |
| Cardiac disorders | n (%) | n (%) | n (%) | n (%) |
| … | n (%) | n (%) | n (%) | n (%) |
| … | n (%) | n (%) | n (%) | n (%) |
| … | n (%) | n (%) | n (%) | n (%) |

The Medical Dictionary for Regulatory Activities (MedDRA) version 22.0 will be used to code all adverse events to a system organ class and a preferred term. The number of participants and subject incidences of adverse events will be summarized for all adverse events, serious adverse events, adverse events leading to withdrawal of investigational product, fatal adverse events and adverse events of interest (e.g., cardiovascular) and will be tabulated by system organ class by visit window.

**Table 5. Adverse events measures by the treatment arm**

| Adverse Event measures | Treatment | | Placebo | |
|---|---|---|---|---|
| | Participant | Incident | Participant | Incident |
| Severity grade | | | | |
| Mild | n (%) | n (%) | n (%) | n (%) |
| Moderate | n (%) | n (%) | n (%) | n (%) |
| Severe | n (%) | n (%) | n (%) | n (%) |
| Life threatening | n (%) | n (%) | n (%) | n (%) |
| Death | n (%) | n (%) | n (%) | n (%) |
| Expectedness | | | | |
| Yes | n (%) | n (%) | n (%) | n (%) |
| No | n (%) | n (%) | n (%) | n (%) |
| Is the event Serious? | | | | |
| Yes | n (%) | n (%) | n (%) | n (%) |
| No | n (%) | n (%) | n (%) | n (%) |
| Relatedness to study intervention | | | | |
| Definitely | n (%) | n (%) | n (%) | n (%) |
| Probably | n (%) | n (%) | n (%) | n (%) |
| Possibly | n (%) | n (%) | n (%) | n (%) |
| Remote | n (%) | n (%) | n (%) | n (%) |
| Not related | n (%) | n (%) | n (%) | n (%) |

#### 3.7 Concomitant Medications

Concomitant medications and medications taken prior to starting study treatment will be summarized by treatment group based on MedDRA System Organ Classes. Medications are considered concomitant if taken during the trial period. Prior medications are medications with the start date and/or end date before study IMP date (randomisation date).

#### 3.8 Report of planned and unplanned unblinding

*Definition of blinding:* Recruiting and consenting clinicians, researchers and senior statistician will all be blind to the allocation. Participants will be associated with a patient information number (PIN) and will not know their allocation. The KCTU randomisation system is linked directly to the IMP management system and pharmacy.

Unblinding will happens for the participant and medical team in case of variceal haemorrhage. If participants have a variceal haemorrhage or their oesophageal varices have increased in grade, they should permanently discontinue the trial IMP, unblinded and standard of care provided, which may include commencement of NSBB and/or offering endoscopic band ligation of oesophageal varices.

The trial Statistician will be fully blind until the Statistical Analysis Plan (SAP) is approved/signed off by the Trial Steering Committee (TSC). After the SAP is approved by the TSC, the trial statistician is planned to become partially blinded and access patient level data coded as A/B. The trial Statistician will then have access to the adherence data and be able to monitor and inform the DMC of the trial adherence of the participants. He will present the closed DMC report to the DMC members.

Throughout the trial, the Chief Investigator (CI) and Senior Statistician will remain fully blind to treatment allocation until after database lock.

After trial completion, at the final combined Trial Steering Committee and Data Monitoring Committee meeting, the Chair of the TSC will be provided with the allocation (intervention and control) after fully interpreting the findings.

#### 3.9 Descriptive statistics for outcome measures

The baseline characteristics (listed in **Table 2**) and will be summarised descriptively for all patients and by trial arm. The primary outcome and secondary outcomes listed in **Table 2** will be summarised at baseline, month 6, 12, 18, 24, 30, 36 timepoints. Each measure will be described overall and by treatment group for complete case data. Means and standard deviation will be presented for normally distributed continuous variables, median, 25$^{th}$ and 75$^{th}$ percentiles for non-normally distributed variables, or frequencies and proportions for categorical variables. We will summarise outcomes, by trial arm, by the time points, and by main demographic classifying variables if appropriate. Descriptive statistics will be provided on noncompliance and dose modifications by treatment group. The EQ-5D data will be summarised descriptively by treatment group and study visit.

##### 3.9.1 Laboratory Test Results

Summary statistics for baseline, six monthly visits by treatment group will be provided for laboratory haematology measures.

##### 3.9.2 Vital Signs

For each treatment and time point of measurement, vital signs measures (systolic and diastolic blood pressure and heart rate) will be summarised using descriptive statistics (mean, SD, median, range), and presented in tabulated form stratified by treatment arm and follow-up timepoint.

### 4 Data analysis plan – Inferential analysis

#### 4.1 Main analysis of treatment differences

##### 4.1.1 Statistical considerations

BOPPP has been designed, and is being implemented, under an intention-to-treat design.

##### 4.1.2 Time of event (survival time)

Definition of Event Time:

The event time in this trial is defined as the duration from the date of randomisation to the occurrence of primary endpoint incidents (time to the first decompensating event of any elements of All-cause decompensation) or secondary endpoint incidents (e.g., variceal haemorrhage and mortality). For patients who do not experience the primary or secondary endpoints, their event time will conclude at the end of the trial (marked by the last patient's last visit).

Censoring Procedure:

For Event-Free Patients at Trial Termination: Patients who are free from events at the trial's conclusion will be censored on the latest date of follow-up. This date will be determined based on the information available in the trial forms or medical notes. For Patients Lost to Follow-Up: Patients who are lost to follow-up during the trial will be reviewed at the study's end using their medical notes. This review will check for any occurrences of all-cause decompensation, variceal haemorrhage, or mortality. Censoring of Patients Without Reported Events: If none of these events are reported in their records, such patients will be considered as part of the event-free population and will be censored on the latest known date when their outcome status is confirmed.

#### 4.2 Analysis Datasets

##### 4.2.1 Intention-to-treat (ITT) population

The intention-to-treat population set will include all participants who randomised, regardless of other outcomes such as compliance and adherence with the assigned treatment regimen, termination of treatment for adverse effects or missed visits.

##### 4.2.2 Per protocol population (PPP)

Definition: The Per Protocol Population set will consist of participants from the Intention-to-Treat (ITT) population who have also adhered to the study protocol. This includes not only receiving the treatment as per the assigned regimen but also complying with all study procedures, most follow-up visits, and any other protocol-specified requirements.

Reason for Exclusion from PPP: Participants who deviate significantly from the protocol (such as poor adherence to the treatment regimen, or other major protocol violations) will be excluded from the PPP set. This exclusion criterion is based on the potential for protocol non-adherence to introduce bias, particularly biases that could affect the assessment of treatment effectiveness.

Analysis Considerations: Analysis of the PPP set is to understand the efficacy of the treatment under ideal conditions where the protocol is followed.

#### 4.3 Data Screening, Cleaning and Acceptance

##### 4.3.1 General Principles

The objective of the data screening and cleaning is to assess the quantity, quality, and statistical characteristics of the data relative to the requirements of the planned analyses.

##### 4.3.2 End of trial - database lock

The primary analysis will be carried out after the end of trial date, and database is locked. The planned end of trial date will be the date when the last randomised participant has been followed-up for up to 36 months or after a minimum of 185 events.[†] After a minimum of 185 events are reported to the DMC, this will trigger a recommendation to the TSC to stop further dispensing of IMP. The final follow up, approximately 7 months later. At this point the final data extract will be requested, dataset will be cleaned. During the data cleaning process all the missing or incomplete data, outliers, and invalid or unacceptable data will be queried.

Database soft lock (database freeze) is the point where all case report form data has been input into the database and all known queries are resolved. When queries are resolved and soft lock is declared, the edit rights of all data entry and data management personnel are revoked so that only the Lead Data Manager (KCTU member) has the ability to modify the database.

At this point final tables, figures and listings will be re-generated. Also, if the clinical study was blinded, the statisticians can now generate programs to unblind the data. Fully Unblinding does not take place prior to hard lock in order to minimise the potential for bias.

Once hard lock is declared, the database is considered final and should not be unlocked. In case of inconsistency, which is associated with a critical data field, the database may need to be unlocked.

The process for unlocking a 'final' database must be pre-defined in the study project and must be supported by the sponsor. Also, when the erroneous data is changed, an audit trail will document what data field was changed, who changed it, when it was changed and why it was changed. The database is then re-locked following the same process as noted previous for the first hard lock and generation of final Tables, Figures, and Listings and the Clinical Study Report are then completed.

---

[†] The trial would be terminated early based on an independent Data Monitoring Committee recommendation and Trial Steering Committee Decision.

For changes to less critical fields, unlocking a 'final' database is not warranted. In this case, a memo is generated to the TMG, and incorporation of the relevant database change can be handled programmatically, with proper documentation.

##### 4.3.3 Data Handling and Electronic Transfer of Data

The King's Clinical Trials Unit (KCTU) will collect, safeguard and extract/provide all data to be used in the planned analyses. This study uses the MACRO-Infermed database. Data Management Plan (DMP) for BOPPP provides a detailed outline how data are to be recorded and handled during the trial.

##### 4.3.4 Survival Data

Survival data will be collected for components of all-cause decompensation, variceal haemorrhage, variceal size progression and mortality (liver related, cardiovascular-related, all-cause). The data comprise the incidence of any of these events and the date of incidence. Patients who do not experience the incidence (even-free patients) will be censored.

In response to the updated primary outcome of time to all-cause decompensation, the study will utilise patient medical notes to track various components of decompensation, ensuring comprehensive follow-up even if a patient misses a scheduled visit. If none of the specified component are not observed, participants will be censored at the date of their last recorded medical note during the trial. This approach allows for a thorough analysis of event-free survival. To minimize missing data, any gaps identified in the extracted dataset during the cleaning process will be addressed through queries. This proactive strategy aims to reduce instances of missing data, thereby enhancing the reliability of the study's findings.

##### 4.3.5 Trial endpoints

**Primary endpoints**

1. Time to first decompensating event.
2. Cost-utility of NSBB over trial follow-up to 3 years.

**Secondary & tertiary endpoints**

1. Estimation of the one, and 3-year variceal haemorrhage rate by allocation, and associated number needed to treat.
2. Progression to medium/large oesophageal varices at gastroscopy requiring clinical intervention.
3. Development of gastric, duodenal, or ectopic or rectal varices in the GI tract at gastroscopy
4. Composite of progression in variceal size and haemorrhage as per 1 and 2 by 3 years.
5. Survival (Overall, liver-related, cardiovascular-related).
6. Quality of life, EQ-5D-5L.

All of the above will be analysed up to 3 years from randomisation. We will seek permission for collection of primary outcome and Health Episode Statistic (HES) data post cessation of IMP until trial completion.

##### 4.3.6 Detection of Bias

Purpose of Randomisation: In the BOPPP trial, randomisation is employed to ensure comparability between treatment groups regarding both known and unknown factors, particularly in a large sample (approximately 740 patients).

Strategies for Detecting Bias:

- Comparing Withdrawal Rates: We will compare the percentage of participants who prematurely withdraw from the treatment in each arm.
- Follow-up Consistency: The rate of missed follow-up appointments for any reason will be compared between the two arms.
- Baseline Characteristic Comparison: A thorough comparison of baseline characteristics, medical history, and severity of medical conditions at baseline will be conducted between the treatment groups. Additionally, stratification factors recorded at baseline will be tabulated to assess any misspecifications during randomisation.
- Medication Adherence and Discontinuation Analysis: The percentages of IMP adherence, IMP discontinuation, and dosage logs will be compared between the treatment groups to identify any discrepancies.
- Site Consistency Check: Consistency in withdrawals and adherence to IMP across different study sites will be examined.
- Adjustment for Imbalance: In cases where there are concerns that key baseline or prognostic factors may not be balanced post-randomisation, resulting in imbalanced groups, covariate adjustment based on these factors may be considered.

##### 4.3.7 Outliers

Potential data outliers will be identified during the screening and cleaning process of dataset and queried as appropriate. Any data points that are identified as possible outliers, but are subsequently verified through the query process, will be treated as valid data, and analysed accordingly.

##### 4.3.8 Statistical Assumptions and Distributional Characteristics

General Approach: For each inferential testing or estimation method used in this study, we will assess the necessary statistical assumptions to ensure validity. This includes examining distributional characteristics of endpoints and residuals.

Cox Proportional Hazards Model Assumptions: The Cox proportional hazards model, a crucial tool in our analysis, relies on several key assumptions, including a) the hazard ratios are constant over time (proportionality of hazards). b) The survival times of different individuals are independent of each other. It's important to check whether these assumptions hold for our data by analysing the residuals of the fitted Cox model. This includes examining the Schoenfeld residuals to assess the proportionality assumption.

Handling Assumption Violations: In cases where the assumptions are not met, these deviations will be adequately described. This may involve transforming endpoint values or employing alternative inferential or estimation methods that better fit the data. The use of transformations or alternative methods will be fully justified and detailed in the final study report. For more information, see section (4.4.5.6).

##### 4.3.9 Statistical Analyses Quality control

Data Cleaning and Analysis Oversight:

- Development and Maintenance: Data cleaning, analysis, and coding will be overseen by both Junior and Senior Statisticians. The Senior Statistician will perform checks and verifications in accordance with current Standard Operating Procedures (SOPs) for quality control.
- Tool for Analysis: The main environment for statistical analyses and output production will be STATA, with the specific version used stated at the time of analysis.

Validation and Consistency Checks:

- Standardized Output Production: Tables, figures, and listings will be generated using validated standard programs to ensure consistency and accuracy.
- Withdrawal and Patient Status Comparison: We will compare withdrawals from the study and patient status to minimize inconsistencies and understand patterns in data dropout.
- Range Checks: Data will be checked for any range constraints, including adherence to eligibility criteria.
- Outcome Date Verification: Dates associated with primary and secondary outcomes will be scrutinized for completeness and consistency.

Additional Quality Control Measures:

- Missing Data Analysis: Investigate patterns of missing data and apply appropriate methods for handling missing data.
- Diagnostic Checks: Perform diagnostic checks on statistical models to ensure the validity of assumptions (e.g., linearity, normality, homoscedasticity).
- Cross-Validation: Use cross-validation techniques, where appropriate, to assess the robustness of the statistical models.
- Audit Trails: Maintain comprehensive audit trails of data manipulation and analysis processes.
- Code Review and Approval: The Senior Statistician (BC) will meticulously review and approve all codes related to data manipulation post-extraction, as well as the statistical analysis itself.
- External Review: Where feasible, engage in external peer review of the analysis plan and key findings for additional validation.
- Documentation of Decisions: Document all decisions made during the analysis process, including rationale for any changes or deviations from the original plan.

#### 4.4 Planned Analyses

##### 4.4.1 General Considerations

• Continuous variables will be summarised by the mean, standard deviation, median, and range. The confidence interval and standard error for the mean will also be summarised for variables that are continuous.

• Categorical variables will be summarised by the number, frequency and percentage in each category.

• Time to event variables will be summarised with hazard ratios, Kaplan-Meier curves, Kaplan-Meier quartiles for important covariates, the number of participants per treatment group, the number of participants censored, and the number & percent of participants with events or at risk.

• Where confidence intervals are provided, these will be 2-sided at the 95% level, unless otherwise specified.

##### 4.4.2 Interim Analysis and Early Stopping Guidelines

No interim analysis is planned for efficacy or harm, beyond the feasibility pilot phase assessment.

###### 4.4.2.1 Internal pilot (Recruitment and retention feasibility)

At 12 months after participant recruitment opens, we will assess feasibility of: recruitment and retention acceptability, with progression criteria:

- At least eight sites opened, with at least one patient randomised at each.
- At least 80 patients randomised.
- At least 70% retention rate.

If we meet the above criteria, we will progress to the end of trial.

##### 4.4.3 Analysis of primary outcome

The primary outcome data will be analysed using the intention-to-treat (ITT) population. Patients who remain event-free will be right-censored as of their last evaluation date, which is typically the end of the trial or the latest recorded evidence in their medical notes.

###### 4.4.3.1 Descriptive Analysis:

Initial descriptive analysis will include hazard ratio exploration and comparative analysis between the two treatment arms (Carvedilol vs placebo). The standard Kaplan-Meier estimate will be used to describe all-cause decompensation.

###### 4.4.3.2 Inferential Analysis for Primary Outcome

The time-to-first event of all-cause decompensation from randomisation will be analysed using a Cox's proportional hazards model. This model will include a random effect for the site, known as a frailty model, to control for unobserved heterogeneity across sites. The proportionality assumptions of the Cox model will be thoroughly assessed, as detailed in section (4.4.5.6). Event times will be recorded in continuous time, with complete dates for events and last monitored dates for right censoring.

Adjustments and Random Effects:

The Cox model will be adjusted for main baseline characteristics, including patient age, gender, disease severity (measured by MELD Score), and cause of liver disease. The inclusion of a random effect for the Site in the model accounts for potential variations between different trial locations. The assumption of proportional hazards will be evaluated using Kaplan-Meier plots, along with at-risk tables for the two groups. Findings will be based on the adjusted hazard ratio (aHR) with associated 95% confidence intervals and p-values.

Sensitivity analyses will be performed as further exploratory analyses for the primary and secondary endpoint (see section 4.6).

##### 4.4.4 Analysis of secondary outcomes

**General Approach:**

Population: Analyses of secondary outcomes will be performed on the Intention-to-Treat (ITT) population.

Random Effects Model: For outcome data that we have clustering by site or participants a generalized linear mixed effect model will be used, with site and participants as a random effect.

**List of secondary outcomes**

- Estimation of the one, and 3-year variceal haemorrhage rate by allocation, and associated number needed to treat.
- Progression to medium/large oesophageal varices at gastroscopy requiring clinical intervention.
- Development of gastric, duodenal, or ectopic or rectal varices in the GI tract at gastroscopy.
- Composite of progression in variceal size and haemorrhage as per 1 and 2 by 3 years.
- Survival (Overall, liver-related, cardiovascular-related).
- Quality of life, EQ-5D-5L.

**Estimation of the one, and 3-year Variceal Haemorrhage Rate:**

Analysis Method: Time-to-event analysis using Cox proportional hazards models. This method will assess the time from randomisation to the first variceal haemorrhage event. The hazard ratio will be calculated, and Kaplan-Meier curves will be used to estimate the haemorrhage rates at 1 and 3 years. Secondary outcomes will be analysed in an equivalent way to the primary outcomes.

**Progression to Medium/Large Oesophageal Varices:**

Analysis Method: Time-to-event analysis using Cox proportional hazards models. This method will assess the time from randomisation to the first occurrence of progression to medium/large oesophageal varices. The model will estimate hazard ratios for this progression, taking into account the treatment arm and adjusted for baseline covariates. Kaplan-Meier curves can be utilised to visualize the time to progression.

**Development of Gastric, Duodenal, Ectopic, or Rectal Varices:**

Analysis Method: Time-to-event analysis using Cox proportional hazards models. This approach will focus on the time from randomisation to the first development of gastric, duodenal, ectopic, or rectal varices. The model will provide hazard ratios associated with these developments, with adjustments for treatment arm and relevant baseline covariates. Kaplan-Meier curves will help in estimating the rates of development at specified time points.

**Composite of Progression in Variceal Size and Haemorrhage:**

Analysis Method: Time-to-event analysis using Cox proportional hazards models. This method will combine the outcomes of variceal size progression and haemorrhage into a single event. The time from randomisation to the occurrence of either event will be analysed. Hazard ratios will be calculated to understand the impact of the treatment arm on this combined outcome, adjusting for baseline covariates. Kaplan-Meier estimates will provide insights into the timing of these events.

**Survival (Overall, Liver-related, Cardiovascular-related):**

Analysis Method: Kaplan-Meier survival analysis and Cox proportional hazards models. These methods will be used to estimate survival rates and hazard ratios for overall, liver-related, and cardiovascular-related mortality.

**Quality of Life, EQ-5D-5L:**

Analysis Method: Linear mixed model regression, including site and participants as random effect in the model, as well as adjusting for baseline covariates.

**Additional Statistical Considerations:**

Adjustment for Covariates: All models will adjust for baseline characteristics like age, gender, cause of liver disease, and disease severity (MELD Score). If appropriate to evaluate the change of these variables or the time, the visit time-points (M6, M12, M18, M24, M30 and M36) will be added as the time-varying covariate to the model.

Random Effects for Site: For outcomes that might vary by study site and are clustered, random effects for the site will be included in the models. When we have more than one observation per participant in any of the models, the subject ID will be added to the model as well as sites to control for the random effect linked to participant.

Model Diagnostics: Model assumptions will be verified, and diagnostics (e.g., checking residuals) will be performed to ensure model fit.

##### 4.4.5 Statistical considerations

###### 4.4.5.1 Compliance to visit time window

Treatment will be initiated immediately after randomisation and dose escalation happens one-week post-randomisation. Outcomes are then measured by researchers every 6 months at months 6, 12, 18, 24, 30 and 36 after randomisation (plus or minus 6 weeks at each interval due to the visit window). We will report the proportion of visits which are outside of the expected visit windows. Gastroscopy outcomes are recorded yearly from baseline (month 12, 24 36) based on the standard of care. We will report the number of participants who fail to start trial IMP after randomisation.

###### 4.4.5.2 Stratification and clustering

Randomisation is stratified by site. Site is included in all Cox's proportional hazards and linear mixed models as a random effect to control for heterogeneity or variability causes by site. If the structure of analysis is longitudinal with repeated measures for outcomes, we will consider a random intercept and slope per cluster (individual) as appropriate.

###### 4.4.5.3 Protocol deviations and violations

A protocol deviation/violation is defined if there is an unplanned excursion from the protocol. A protocol deviation (PD) is defined as a non-serious breach from the protocol that is unlikely to lead to any impact on the value of the data contributing to the overall treatment effect. A protocol violation (PV) is an excursion from the protocol that is more serious and likely to lead to a meaningful impact on the quality of the data and would lead the patient(s) from being excluded from the per protocol population. The list and frequency of PD and PV will be tabulated per treatment arm. Non-adherence or non-compliance are considered as a form of protocol deviation.

###### 4.4.5.4 Method for handling multiple comparisons

There is one primary outcome and we only comparing two groups. We will not adjust for multiple comparisons.

###### 4.4.5.5 Method for handling non-compliance

In addition to the primary ITT analysis, the effect of actually receiving treatment as defined in the protocol will also be estimated. We will analyse primary outcomes for a per protocol subset of the data. This will exclude individuals who are not compliant (less than 50%).

###### 4.4.5.6 Model assumption checks

**Checking Normality in Models:**

Normality Assumption: For models assuming normal distribution of residuals or covariates, normality will be checked using plots and statistical tests. In cases of significant deviation from normality, data transformations (such as log, square root, etc.) will be considered to achieve normality.

**Proportional Hazards Assumption in Cox Regression:**

The Cox proportional hazards model assumes that the hazard ratios between any two groups are constant over time. Kaplan-Meier survival curves and log-log [-log(-log(survival probability))] plots against log(time) will be used to visually inspect the proportionality assumption. Crossing curves or divergent trends suggest a violation of this assumption. Schoenfeld residuals will be used to statistically test the proportionality assumption. This involves correlating the residuals with time to check for any time-dependent patterns.

Dealing with Non-Proportionality: If evidence of non-proportionality is found, several approaches may be employed when appropriate.

- Extended Cox Model: Include time-dependent covariates in the Cox model to allow hazard ratios to change over time.
- Alternative Models: Consider using alternative models such as the proportional odds model or accelerated failure time models.
- Stratification: Stratify on variables causing non-proportionality if they are not of primary interest.

Nonlinear Covariate Relationships:

The assumption of linearity in the Cox model for continuous covariates will be assessed using residual plots. Plotting residuals or partial residuals against continuous covariates will help detect nonlinearity. If nonlinearity is detected (evident from patterns or curves in the residual plots), we will consider using transformations of the covariates or including polynomial or spline terms to model the non-linear relationship adequately.

**Lack of independence:**

Because BOPPP is a multi-site trial, we assume such design feature likely to produce correlated outcomes within sites or heterogeneity between sites. We will implement frailty models in the analyses which account for correlation within a group by introducing a random effect.

**Assumption check for Linear Mixed Models (LMM)**

For continues secondary variables when using the LMM we will check the linearity and homogeneity (check by plotting residuals vs predicted values), normality of error term (check by histogram, Q-Q plot of residuals, Kolmogorov-Smirnov test).

#### 4.5 Estimand Framework

##### 4.5.1 Definition of Primary Estimand

The primary clinical question of interest for this trial is: What is the difference in the time to first all-cause decompensating event in patients with cirrhosis and small oesophageal varices (as defined by the trial's inclusion/exclusion criteria), over a maximum of 36 months follow-up, treated with Carvedilol compared to Placebo.

The primary estimand for this trial is described by the following attributes:

##### 4.5.2 Target Population:

Patients diagnosed with cirrhosis, specifically those identified with small oesophageal varices (≤5mm in diameter) and without prior variceal haemorrhage.

##### 4.5.3 Variable (or Endpoint):

Time to the first occurrence of any all-cause decompensating event (including variceal haemorrhage, new or worsening ascites, new or worsening hepatic encephalopathy, spontaneous bacterial peritonitis, hepatorenal syndrome, a significant increase in Child Pugh Grade (increase by one grade) or MELD score (increase by 5 point), and liver-related mortality).

##### 4.5.4 Treatment or Intervention:

Carvedilol, initiated at a dose of 6.25 mg once daily, with an up-titration to 12.5 mg daily based on tolerability and target heart rate, after one week and continuing for three years.

##### 4.5.5 Population-Level Summary:

The adjusted hazard ratio (aHR) of the first all-cause decompensating event between the Carvedilol and placebo groups over the 3-year follow-up period.

##### 4.5.6 Post-Randomisation Events (Intercurrent Events):

###### 4.5.6.1 Discontinuation of the IMP

Intercurrent events which can lead to the discontinuation of the intervention medication (Carvedilol or Placebo), are primarily related to liver disease progression and treatment-related issues. They include:

**Disease progression (Progression in Varices):**

- Progression to medium or large oesophageal varices.
- Development of gastric, rectal, or ectopic varices as observed in gastroscopy.

For patients experiencing these events, six-monthly follow-up data collection will continue to monitor disease progression and treatment outcomes.

**Disease progression (Progression-Related Interventions):**

- The need for a Transjugular Intrahepatic Porto-Systemic Shunt (TIPS).
- Liver transplantation.

Following these interventions, routine six-monthly data collection will not be continued.

**Clinician directed permanent discontinuation from IMP:**

- Discontinuation directed by clinicians, which can occur due to adverse events (AEs) or serious adverse events (SAEs).

For patients who permanently discontinue the IMP for these reasons, six-monthly follow-up data collection will still be pursued.

In all the above scenarios, despite the discontinuation of the IMP, efforts will be made to collect vital data until the end of the study. This includes mortality, variceal haemorrhage, and Health Episode Statistics (HES) data. The aim is to minimise the impact of missing data and to ensure robustness in the trial's outcome analysis.

It is highly probable that there will be a recorded increase in the MELD score or Child-Pugh score before any intercurrent events listed above as disease progression. In such cases, the disease progressions mentioned in the intercurrent events will not influence the primary outcome estimation.

##### 4.5.7 Handling of Intercurrent Events in Analysis:

###### 4.5.7.1 Treatment Policy Strategy:

For the primary analysis, a treatment policy strategy will be employed. This means that occurrences of intercurrent events like progression in varices, progression-related interventions (TIPS, transplant), and permanent discontinuation from IMP due to clinician direction will be considered irrelevant in defining the treatment effect. The analysis will use the values for the variable of interest regardless of whether these intercurrent events occur.

###### 4.5.7.2 Supplementary Analyses:

If the rates of occurrence of these intercurrent events are non-trivial, separate supplementary analyses will be conducted. These analyses will explore different handling strategies for the intercurrent events to assess their impact on the treatment effect.

###### 4.5.7.3 Secondary Outcomes:

The secondary outcomes will also follow the treatment policy strategy for handling intercurrent events.

###### 4.5.7.4 Exploratory Analyses:

Depending on the data, exploratory analyses might be carried out using different strategies like the hypothetical scenario strategy or the composite variable strategy. These strategies can provide additional insights into the impact of intercurrent events on the overall treatment effect.

###### 4.5.7.5 Sensitivity Analyses:

Sensitivity analyses will be conducted to assess the robustness of the primary analysis results. These analyses will test different assumptions about the handling of intercurrent events and their impact on the treatment effect.

#### 4.6 Sensitivity analyses

Sensitivity Analysis is defined as “a method to determine the robustness of an assessment by examining the extent to which results are affected by changes in methods, models, values of unmeasured variables, or assumptions”. (Schneeweiss, 2006) Robustness refers to “the sensitivity of the overall conclusions to various limitations of the data, assumptions, and analytic approaches to data analysis”.

We will perform sensitivity analyses to check if missing data, non-compliers, protocol deviators/violators, and intercurrent events will change the overall conclusion of the trial observed results. If, after performing sensitivity analyses the findings are consistent with those from the primary analysis and would lead to similar conclusions about treatment effect, the researcher is reassured that the underlying factor(s) had little or no influence or impact on the primary conclusions. In this situation, the results or the conclusions are said to be “robust”.

For sensitivity analyses we assess the robustness of the results of ITT to protocol deviations (per-protocol analysis: in which participants who violate the protocol are excluded from the analysis). The per-protocol analysis provides the ideal scenario in which all the participants comply and is more likely to show an effect; whereas the ITT analysis provides a “real life” scenario, in which some participants do not comply.

##### 4.6.1 Planned subgroup analyses

Particularly, in the condition that the overall intervention effect is present, analysis of the results by appropriate subgroups may help to identify the specific population most likely to benefit from the intervention, or the consistency of the benefit with respect to the primary efficacy endpoint of all-cause decompensation and secondary endpoint such as variceal Haemorrhage or size progression. Subgroup analysis may also elucidate the mechanism of action of the intervention. Selection of subgroups are only relying on baseline data, and not data measured after the initiation of the intervention.

Analysis and plots for subgroups will be provided for the following some of the variables such as: age category, sex, main geographical region, ethnic groups, cause of liver disease (Alcohol related, NALFD, viral hepatitis, autoimmune), severity of the disease (Child Pugh Score: A vs B or MELD Score), and ascites (yes/no).

The Child Pugh Score will be dichotomised to Grade A (5-6 points) vs Grade B (7-9 points). The MELD Score will be classified to low vs high binary codes. The cut off threshold will be determined at TSC meeting.
We will not emphasis on the subgroup analyses for variables which the study is not powered to detect treatment effects for them.
Any conclusions drawn from subgroup hypotheses not explicitly stated in the protocol will be given less credibility than those hypotheses stated a prior.

#### 4.7 Exploratory analyses

None planned beyond the descriptive analysis.

#### 4.8 Exploratory mediator and moderator analysis

No mediation or moderation analysis is planned as part of the primary publication.

### 5 Software

#### 5.1 Data management

An online data collection system for clinical trials (MACRO: InferMed Ltd) will be used. This is hosted on a dedicated server at KCL and managed by the King's Clinical Trials Unit (KCTU).

#### 5.2 Statistical analysis:

Stata (version 18 or later) will be used for data cleaning, description, and the main inferential analysis. For any statistical analysis report reference will be made to the trial protocol and SAP number.
